# Brain and physiological responses to flavored waters with different sweeteners: a randomized cross-over study in healthy young adults

**DOI:** 10.64898/2025.12.08.25341825

**Authors:** Paul A.M. Smeets, Ralf Veit, Els Oosterink, Saskia Meijboom, Davide Risso, Hubert Preissl, Stephanie Kullmann

**Affiliations:** Division of Human Nutrition and Health, Wageningen University & Research, Wageningen, The Netherlands; Institute for Diabetes Research and Metabolic Diseases of the Helmholtz Center Munich at the University of Tübingen, Tübingen, Germany; German Center for Diabetes Research (DZD), Tübingen, Germany; Internal Medicine IV, Department of Diabetology, Endocrinology and Nephrology, Eberhard Karls University Tübingen, Tübingen, Germany; Institute of Pharmaceutical Sciences, Department of Pharmacy and Biochemistry; Interfaculty Centre for Pharmacogenomics and Pharma Research at the Eberhard Karls University Tübingen, Tübingen, Germany; Wageningen Food and Biobased Research, Wageningen University & Research, Wageningen, The Netherlands; Tate & Lyle PLC, London, UK

**Author notes:** Correspondence: Paul Smeets.

## Abstract

**Background:** Drinks with low-no-calorie sweeteners (LNCS) do not contribute to energy intake while still providing a hedonic experience through sweetness. LNCS can have differential effects on brain areas involved in food intake and reward compared to sugars.

**Objective:** To determine changes in brain activity and the effect on physiological markers following the ingestion of flavored waters sweetened with the sugar sucrose or LNCS.

**Methods:** 30 healthy individuals participated in a randomized crossover study with six treatments. Participants were scanned after an overnight fast using magnetic resonance imaging, including arterial spin labelling to measure cerebral blood flow (CBF), before and after ingestion of 500-ml drinks: Water, or equi-sweet flavored waters with 25 g sucrose, sucralose, stevia extract, allulose+stevia extract or monk fruit extract. CBF was measured at baseline, 5 and 30 min; gastric content volume at baseline, 25 and 45 min. Serum insulin and glucose were measured and participants rated their appetite and thirst throughout each visit.

**Results:** Hypothalamus CBF was not differentially affected by any of the drinks. In the ventral tegmental area (midbrain), treatment effects differed, with lower CBF after sucrose compared to water, sucralose and monk fruit drink ingestion at 30 min. Exploratory whole-brain analyses showed differential CBF changes for the allulose+stevia (amygdala) and stevia (putamen) drinks compared to sucrose. Despite its low energy content, the allulose+stevia drink delayed gastric emptying similar to the sucrose drink, while only sucrose increased glucose and insulin levels.

**Conclusions:** Although flavored waters with LNCS mostly elicit similar neural and gastrointenstinal responses as water, they have some distinct effects on the brain compared with 25 g of sucrose, particularly in reward-related brain areas. Further exploration of the neural and physiological effects of allulose and stevia and dose-dependent investigations are warranted.

## Introduction

The brain plays a crucial role in regulating energy homeostasis and eating behavior. Because of the global obesity epidemic and associated pathology there is great interest in strategies to reduce energy intake. One very common strategy is to replace sugar with low-no-calorie sweeteners (LNCS). This strategy is prominently applied in soft drinks because sugar-sweetened soft drinks have been shown to contribute to overconsumption and obesity^1,2^. There has been much controversy about potentially undesirable effects of providing sweetness, with or without calories, although empirical evidence does not support that exposure to sweetness stimulates a greater liking and desire for sweetness and greater intake of sweet foods^3,4^. Though they may sustain sweetness preference if used frequently^5,6^. Similarly, a wealth of human evidence suggests that overall non-nutritive sweetened drinks are equivalent to water, in terms of their acute metabolic effects and their effects on energy intake and weight^7,8^. However, at the same time there is mounting evidence that not all LNCS have the same physiological effects because they are processed differently in the body^9–11^, as is also the case for different types of sugar which can have different glycemic responses^12^. Differences among (non-nutritive) sweeteners start already with differences in their sensory properties^13,14^ and are also apparent in their metabolic pathways and effects on the gut microbiota^15,16^. Interestingly, it has been shown that blocking glucose taste sensation decreases the speed of gastric emptying as well as glycemic metabolism both during and after glucose ingestion^17^. However, this has not been observed for the sweetener aspartame^18^.

Several brain imaging studies using functional magnetic resonance imaging (fMRI) have sought to examine the brain responses to simple solutions or drinks with sugars, LNCS or only calories (non-sweet carbohydrates; maltodextrin)^19–22^. Although some of these studies point to a difference between sugar and LNCS the results are not very consistent. One potential limitation of such taste fMRI studies is the repeated administration of numerous very small sips over an extended period, which does not reflect naturalistic consumption patterns. Examining the response to ingestion of a regular consumed amount (200-500 ml) is more similar to real-life situations and thus more ecologically relevant. Most of the studies employing the latter approach have used blood oxygen level-dependent (BOLD) fMRI, the same MRI measure that is used in taste cue fMRI studies. Their most consistent finding is a decrease in the BOLD signal particularly in the hypothalamus after ingestion of 75 g sugar but not water^23–28^. A decrease in hypothalamic activity has been considered a biomarker for satiety, reflecting the reduced drive to eat after nutrient intake^29^. Conversely, an increase in hypothalamic activity in response to fructose has been observed in some studies and is thought to reflect incomplete appetite suppression^29^.

However, with respect to different LNCS, there are very few studies and these do not yet provide consensus with regard to consumption-induced BOLD signal changes as well as changes in resting state functional connectivity^27,30^. Meyer-Gerspach et al. used arterial spin labeling (ASL) to quantify cerebral blood flow (CBF), next to resting state fMRI (rsfMRI), to compare the effects of 300-ml intragastric loads of xylitol, erythritol, glucose and water^31^. They found that xylitol, but not erythritol, increased CBF in the hypothalamus, whereas 75 g glucose had the opposite effect. In line with other studies comparing different sweeteners, changes in resting state functional connectivity patterns showed both similarities and differences in brain network properties following xylitol, erythritol, and glucose administration^31^.

A recent study in young adults with varying body weights showed that, among those with a healthy weight, a sucrose-sweetened drink decreased lateral hypothalamus blood flow more than a sucralose-sweetened drink, but not more than water^32^. In addition, they found that sucralose, compared to sucrose and water, increased resting state functional connectivity between the hypothalamus and brain regions involved in motivation and somatosensory processing. This adds to the notion that different kinds of sweeteners can have differential effects on the brain although the implications of this e.g. with regard to appetite regulation are not very clear yet.

Beverages are among the most important sources of added sugar and caloric intake^33,34^. Therefore, LNCS are widely used to prevent increased energy intake from beverages, while still providing the hedonic experience of sweet taste. Previous studies have used different sweetener loads, usually administered as simple solutions in water that are ingested or infused. Next to classical carbonated soft drinks, flavored waters have been introduced as less sweet alternative sweet beverages. They may constitute an easier-to-drink and more palatable and ecologically relevant alternative which maintains the relative simplicity of simple solutions but is available on the consumer market in different flavors and with different sweetening agents, both caloric and low-no-calorie. The global and in particular the European market for flavored waters is growing^35^, especially among Generation Z/Millennials.

In the current study we aimed to determine the changes in regional brain activity, blood glucose and insulin levels, and gastric emptying in response to the ingestion of different flavored waters in individuals with healthy body weight. Beverages were sweetened with either the nutritive sugar sucrose, the LNCS sucralose, stevia extract, monk fruit extract, or the low-calorie sugar replacement allulose (in combination with stevia extract). *A priori* regions of interest included the hypothalamus, which is a key homeostatic region sensitive to energy intake, and brain reward-related regions such as the ventral striatum and midbrain which include core dopaminergic areas responsive to food cues and energy intake (nucleus accumbens (Nacc), ventral tegmental area (VTA), substantia nigra (SN)). In addition to testing several novel sweeteners in a consumer-available type of sweet beverage-some of which (i.e., allulose and stevia in combination, monk fruit) have not been investigated previously in this context-this comprehensive study also leveraged the advantage of measuring cerebral blood flow (semi-quantitatively) alongside physiological responses, rather than assessing only a subset of these measures. The overall hypothesis was that, while the sugar sucrose will elicit a response in brain areas related to food intake regulation and reward, equisweet low or non-nutritive sweeteners might not generate a similar response due to the lack of, or lower, energy content in individuals with a healthy body weight.

## Methods

### Study design

This study was a double-blind randomized crossover trial in which healthy participants underwent brain imaging (fMRI), gastric MRI and blood sampling at baseline and after consumption of water or a sweet flavored water. Participants, investigators and statisticians were all blinded to the treatment allocation until data analysis was completed. The primary outcome was CBF in *a priori* defined homeostatic and reward-related brain regions of interest (ROIs; hypothalamus, ventral striatum, dopaminergic midbrain). Secondary outcomes were seed-based functional connectivity (resting-state fMRI), plasma glucose and insulin and gastric content volume. Functional connectivity results will be reported elsewhere.

In addition, subjective ratings of hunger, fullness, desire to eat, thirst, bloating and wellbeing were collected. The study procedures were approved by an accredited Medical Research Ethics Committee (MREC Oost-Nederland, NL81742.091.22) in accordance with the Helsinki Declaration of 1975 as revised in 2013. The study was registered with clinicaltrials.gov under number NCT05575687 in October 2022. All participants signed informed consent.

### Sample size estimation

The hypothalamus was the main *a priori* region of interest. Many fMRI studies have shown that the BOLD signal in the hypothalamus decreases after the ingestion of sugars, but not water^24,25,27,36,37^. In addition, there was one previous fMRI study that employed a similar intervention and primary outcome measurement (CBF) as used here^31^. In line with the BOLD fMRI studies done, this study observed a lower regional CBF after ingestion of 75 g of glucose, compared with water. The average difference was ∼2.4 ml/100g/min. It is also known that the decrease in hypothalamic activity after glucose ingestion is dose-dependent^36^. Accordingly, in the current study we wanted to be able to detect a difference in the hypothalamus between the 25-g sugar-containing drink and water i.e., a difference of 2.4/3 = 0.8 ml/100g/min. The standard deviation was expected to be of similar magnitude. To establish whether treatment effects differ from those in the water control condition, 5 treatments would be compared with water or sucrose, as the primary analysis to show any sweetener’s effect. To account for multiple testing we apply a Bonferroni correction and thus use an alpha of 0.05/5 = 0.01. Using the calculator for crossover studies at http://hedwig.mgh.harvard.edu/sample_size/js/js_crossover_quant.html, we estimated that n=30 datasets would be sufficient for 85% power of detecting a lower CBF in the hypothalamus. We also conducted a more advanced sample size estimation using the GLIMMPSE online tool for mixed models at https://samplesizeshop.org/glimmpse-power-software/. This estimation showed that n=30 would be sufficient to detect a time x treatment interaction effect using a corrected threshold of P=0.01, with powers ranging from 81 to of 91% depending on the assumptions regarding mean effect size and variability (set as above, and plus and minus 20%) and the type of test used (Hotelling Lawley Trace or Greenhouse-Geisser corrected). The within-treatment change in activity from baseline was expected to be larger than the difference between treatments^27^. Therefore, this sample size would also allow us to observe within-treatment changes in regional CBF, even if these are smaller than seen after sucrose, which was expected to have the strongest effect.

### Participants

We recruited healthy (self-reported), right-handed individuals aged 18–30 years with a BMI between 18.5 and 25 kg/m^2^, a sufficient blood hemoglobin level (women > 7,5; men > 8.5 g/dl) and having antecubital veins suitable for blood sampling via a catheter. Exclusion criteria were: Having disturbances of glucose metabolism such as being prediabetic or diabetic; Use of medication that could influence study results including insulin/metformin/proton pump inhibitors, antacids, anti-depressants; Allergy or intolerance for any of the study products/compounds (sucrose, sucralose, stevia extract, allulose, monk fruit extract); Being a regular smoker (smoking more than one cigarette or e-cigarette with nicotine per day); Drinking more than 14 glasses of alcohol a week; Having genetic, psychiatric or neurological diseases affecting the brain; Having a gastric disorder or regular gastric complaints such as heart burn (more than once per week); Having a renal or hepatic disease; Using recreational drugs more than once per week (e.g. marihuana, laughing gas); Having given a blood donation in the past two months; Being pregnant, lactating or planning on becoming pregnant during the study; Currently following or having followed a calorie-restricted diet in the past two months; Having a contra-indication to MRI scanning (including but not limited to pacemakers and defibrillators, ferromagnetic implants, and claustrophobia).

Participants were recruited between January and December 2023 via digital advertisements (e-mail and social media) and invited for an online information session where they could ask questions. Those that appeared eligible and were willing to participate where invited for a screening session. During this session they signed informed consent, filled in the inclusion questionnaire and practiced drinking in a supine position in a mock MRI scanner by consuming 250 ml water through a tube, exactly as in the test sessions. N=35 were included in the study after screening. Of the 35 those, n=2 were unresponsive and never scheduled for a test session, n=2 resigned after two test sessions and n=1 resigned after one test session. N=30 participants (15 females/15 males, age 22.7±2.3 years, BMI 22.2±1.7 kg/m^2^) completed six test sessions (see **Supplementary Figure 1**).

### Treatments

The treatment consisted of the ingestion of 500 ml water or one of five equisweet lemon-lime flavored waters sweetened with sucrose (25 g, 97 kcal), sucralose, stevia extract, monk fruit extract or allulose + stevia extract. This amount is realistic for this beverage category and has been shown to elicit a detectable glycemic response at 30 minutes^38,39^. The water control was not flavored because the flavor could not be matched to that of the flavored waters due to the absence of sweetness. Allulose (25 g in 500 ml, 5%, 5 – 10 kcal) was used in combination with stevia extract following the rationale that 1) in commercial application allulose does not usually fully replace sucrose in beverages but is used in combination with other sweeteners, stevia extracts in particular, to reach higher sugar reduction levels, improve taste quality and reduce overall sweetener costs, and 2) this dose will safeguard gastrointestinal tolerance: allulose is considered safe for human consumption up to 24–36 g (0.4 – 0.6 g/kg/day for a 60 kg individual) when consumed in one sitting, with a recommended maximal single dose where no gastrointestinal side effects are observed of 25 g (FDA, n.d.; Han et al., 2018).

The drinks were made by the Tate & Lyle Innovation Centre (Lübeck, Germany) and provided in 500-ml pasteurized servings in plastic bottles labeled with a letter (A-F) for blinding. All drinks looked the same, like water (clear liquid). Detailed composition can be found in **Supplementary Table 1**. Treatment orders were determined by a research assistant by randomly sampling 6 sequential numbers (representing the six drinks) 40 times with the base function *sample* in R, including the *prob* (i.e., probability) argument to enforce a balanced distribution.

The drinks were consumed at room temperature and participants were instructed to finish within 5 minutes, which they all did. The flavored drinks are briefly referred to by the sweetener used, capitalizing the first letter e.g. Sucrose or Stevia.

### Study procedures

Study procedures are summarized in **Figure 1**. Subjects were instructed to consume the same evening meal before each visit, to refrain from alcohol consumption that evening, and to fast from 10 p.m. onwards (no food or drinks except water). Drinking water was allowed up to 90 min prior to their visit. Upon arrival at Hospital Gelderse Vallei (Ede, The Netherlands), a cannula was placed in an antecubital vein and a blood sample was taken in case the treatment was a sweet drink. Subsequently, baseline MRI scans were performed and verbal subjective ratings were obtained regarding hunger, fullness, desire to eat, bloating, wellbeing and thirst (100-unit scale with 0 = not at all and 100 = very much). After this, participants consumed one of the drinks in a supine position through a drinking tube (similar to drinking through a straw). After the first sip they verbally scored liking and sweetness on a similar 100-unit scale as the other ratings. All participants finished their drinks within 5 minutes.

**Figure 1.**
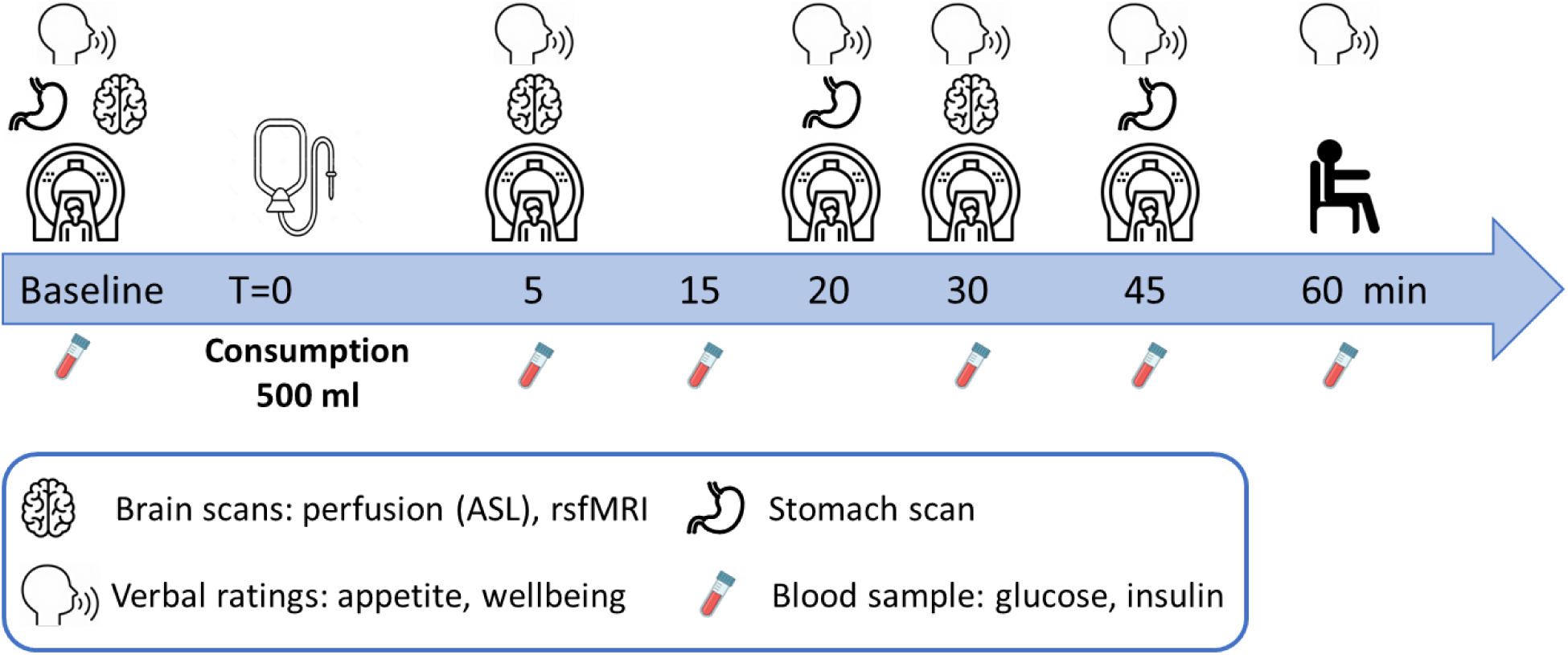
Overview of a test day. For the water treatment there was no blood sampling. T=0 is the start of consumption.

After drinking the T=5 perfusion scan was made, followed by the resting state fMRI scan and the T=20 stomach scan. At T=30 the brain scans were repeated followed by another stomach scan at T=45. After this the participant was taken out of the scanner and seated. Except for water treatment days, post-consumption blood samples were taken at T = 5, 15, 30, 45 and 60 min. Appetite and wellbeing ratings were collected at T=20, 30, 45 and 60 min. After this, participants were offered a takeaway breakfast.

### MRI scanning

Participants were scanned head-first in a supine position with the use of a 3 Tesla Philips Ingenia Elition X MRI scanner (Philips, Eindhoven, The Netherlands) equipped with a dStream torso coil and a 32-channel head coil. The following scans were made:

Stomach scan (18 s): a 2-D Turbo Spin Echo sequence (33 5-mm slices, 1.4 mm gap, acquired in-plane voxel size 1.8 x 1.8 mm, reconstructed to 1 × 1 mm, repetition time 550 ms, echo time 80 ms, flip angle 90°, field of view 400 x 336 x 210 mm, SENSE factor anterior-posterior 2.5) was used with breath hold command on expiration to fixate the position of the diaphragm and the stomach.

Anatomical brain scan (4 min), obtained at baseline: A high-resolution T_1_-weighted 3D fast field echo anatomical scan (repetition time = 9.8 ms, echo time = 4.5 ms, flip angle = 8°, field of view = 256 × 243 × 180 mm, 450 sagittal slices, scanning voxel size = 0.8 × 0.8 × 0.8 mm, reconstruction voxel size 0.4 × 0.4 × 0.4 mm).

Perfusion scan (5 min 20 s): To determine regional CBF, Pseudo-Continuous Arterial Spin Labeling (pCASL, see Alsop et al.^42^) was performed with a segmented 3D readout using GRASE^43^. In total, nine pairs of label and control images were acquired in anterior-posterior phase encoding (PE) direction including two M0 images (labeling distance 90 mm, post-label delay 1800 ms, repetition time = 4148 ms, echo time = 12 ms, flip angle = 90°, field of view = 240 × 240 × 102 mm, 17 axial slices, acquired voxel size 3.74 x 4 x 6 mm, reconstructed voxel size 3 x 3 x 6 mm). After each measurement, two additional M0 images were acquired with reversed PE direction (Posterior-Anterior).

### MRI image analysis

#### CBF

First, images were converted from DICOM to Nifti using dcm2niiX version v1.0.20020720. The T_1_-weighted images were used for segmentation, bias correction and normalization of the anatomy using fsl_anat. The ASL data were analyzed using FSL 6.0.6.4 and BASIL tools. First, motion correction was performed for each series (mcflirt). Thereafter distortion correction was carried out with the M0 images in different PE directions and the correction was applied to the ASL images (topup). For calibration into absolute physiological units, voxel-wise mode was selected. Moreover, we chose ‘Adaptive spatial regularization on performance’ to reduce noise in the CBF map. Finally, partial volume correction was applied to restrict the analyses to gray matter and the images were smoothed with an isotropic Gaussian kernel (full width at half maximum: 6 mm). For ROI analyses we used the flag –region-analysis that allowed extraction of mean CBF values from regions taken from the Harvard-Oxford cortical and subcortical atlases. Mean CBF values were extracted from homeostatic and reward-related brain areas of interest (**Supplementary Figure 3**): the hypothalamus, Nacc (ventral striatum), and VTA and SN (dopaminergic midbrain). The hypothalamus ROI was based on the Neudorfer atlas^44^. The Nacc was based on the Harvard-Oxford Subcortical Structural Atlas as implemented in the FSL software. The VTA ROI was based on the probabilistic VTA atlas of Trutti et al.^45^ accessible in the OpenScience Framework at https://osf.io/9pzj3. The SN ROI was taken from the automated anatomical labeling atlas 3^46^ available at http://www.gin.cnrs.fr/en/tools/aal/. The ROIs are ilFor all ROIs CBF values were normalized by expressing them as a percentage of the global CBF ((ROI CBF/global CBF)*100).

#### Stomach scan image analysis

Total gastric content volume was determined for each stomach scan with the help of an in-house software tool that performs a deep learning-based segmentation of the stomach contents based on an nnU-Net framework^47^. The resulting mask images were checked, and where necessary manually corrected, with the use of the ITK-SNAP program^48^ (http://www.itksnap.org/). An example of a stomach scan time series for all treatments is shown in **Supplementary Figure 5**.

### Blood sample collection and analysis

Blood samples were drawn from the cannula into sodium-fluoride and lithium heparin tubes. After collection, all tubes were centrifuged at 1300 *g* for 10 min at 20 °C, to obtain blood plasma. Blood plasma aliquots were stored at –80 °C until they were analyzed in bulk by a clinical chemistry laboratory (Ziekenhuis Gelderse Vallei, Ede, The Netherlands). To determine glucose concentration, the sodium-fluoride plasma samples were processed using the Atellica CH Glucose Hexokinase_3 (GluH_3) assay kit and quantified using the Atellica CH analyzer (Siemens Healthineers, Netherlands). The lower detection limit was 0.2 mmol/l and the maximum intra-assay CV was 4.5 %. The lithium heparin plasma samples were processed and insulin was quantified using a solid-phase, enzyme-labeled chemiluminescent immunometric assay (IMMULITE 1000 Insulin). The detection range was 2 – 300 μIU/l and inter-assay CVs ranged from 3.3 to 19.1 %. Values below the detection limit were set at 1 μIU/l.

### Statistical analysis

Statistical analyses were performed using RStudio (version 4.4.1, 2024.04.0) and the packages dplyr (1.1.4), emmeans (1.10.3), ggeffects (1.7.0), multcomp (1.4-25) and nlme (3.1-164) on data from the 30 participants that completed six test sessions. Missing values were retained and not imputed. Primary analyses were performed blinded for treatment. For water, blinding was not complete because no blood samples were taken at water visits and it was the only non-sweet drink. Differences in glucose, insulin, gastric content volume, ROI CBF and subjective ratings over all time points were tested using linear mixed models (lme), with the treatment*time interaction as a fixed factor, baseline values as a covariate and participant as a random factor (random intercept). For the ROI CBF and gastric content volume analyses, sex was added as an additional covariate because sex can affect brain function^49^ including perfusion^50^ and digestion^51^. False discovery rate-corrected post-hoc t-tests were used to compare treatments and time points.

In addition, we performed an exploratory whole-brain analysis in SPM12. For this, the CBF maps were entered into a 3 x 6 flexible factorial model with a subject factor and timepoint and treatment as within subject factors. Global CBF was added as a covariate. The sex covariate was omitted because there were no sex by treatment interactions in the ROI analysis. Moreover, the design structure, while appropriate for modeling repeated measures, constrains the inclusion of covariates to those that vary within subjects (i.e., time-varying covariates). Sex is constant across conditions and subjects and is treated as a between-subject covariate that would effectively be orthogonal to the modeled within-subject effects. We created contrasts for the main effect of time (i.e., collapsed over all conditions) between T5 and baseline as well as between T30 and baseline. Moreover, we computed interactions between sucrose as a reference condition and the other conditions separately for baseline vs T5, baseline vs T30 and T5 vs T30. A primary statistical threshold of P < 0.001 uncorrected and a P < 0.05 family-wise error (FWE) corrected for multiple comparisons at the cluster level was applied. Small volume correction (SVC) was applied for the *a priori* subcortical ROIs (hypothalamus, midbrain (both VTA and substantia nigra), the striatum (both ventral and dorsal) and the amygdala.

## Results

### Drink perception

Data are shown in **Supplementary Figure 2**. Liking did not differ among the sweet drinks. All drinks except Monk fruit were liked significantly more than water (all p<0.05). Perceived sweetness of all drinks was greater than that of water (all p<0.001). Sweetness did not differ significantly among the flavored drinks (all P>0.1, Tukey’s HSD post hoc test).

### Cerebral blood flow response in ROIs

Drink-induced changes in CBF in *a priori* homeostatic and reward-related brain regions of interest were assessed (**Table 1**). For the hypothalamus, the primary ROI, there was no effect of timepoint (P=0.26) and treatment (P=0.39), and no interaction between timepoint and treatment (P=0.68). For the nucleus accumbens (Nacc, located in the ventral striatum), there was no treatment effect (P=0.33), but an effect of timepoint (P=0.006), and no interaction between timepoint and treatment (P=0.27). Across all treatments, CBF in the Nacc was greater at T=30 than at baseline (difference 2.61±0.81, P_FDR_=0.004).

**Table 1.**
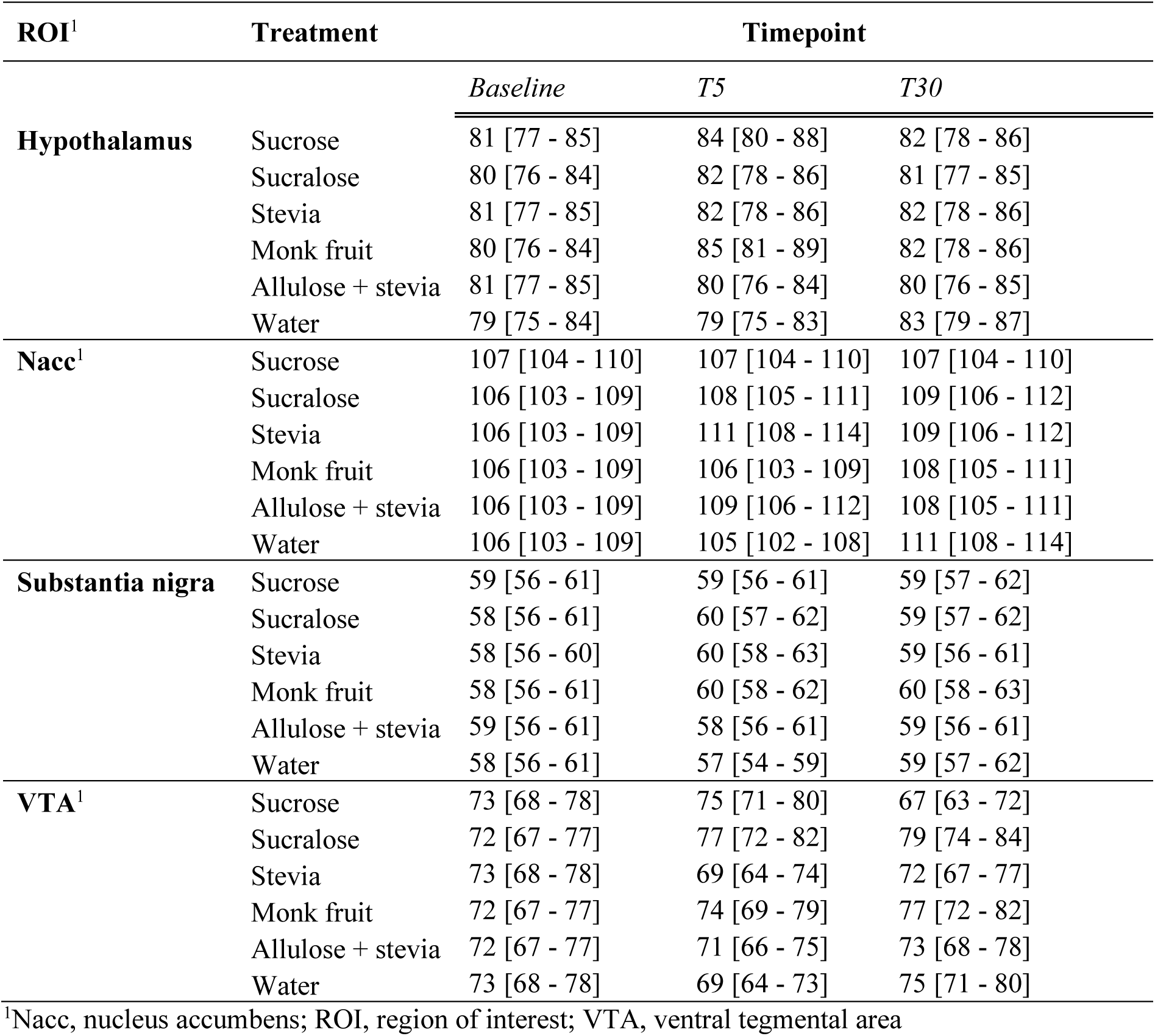
Estimated marginal mean normalized CBF values (% of global CBF) and 95% CI for the ROIs, adjusted for baseline and sex.

| ROI<sup>1</sup> | Treatment | Timepoint |  |  |
| --- | --- | --- | --- | --- |
|  |  | *Baseline* | *T5* | *T30* |
| **Hypothalamus** | Sucrose | 81 [77 - 85] | 84 [80 - 88] | 82 [78 - 86] |
|  | Sucralose | 80 [76 - 84] | 82 [78 - 86] | 81 [77 - 85] |
|  | Stevia | 81 [77 - 85] | 82 [78 - 86] | 82 [78 - 86] |
|  | Monk fruit | 80 [76 - 84] | 85 [81 - 89] | 82 [78 - 86] |
|  | Allulose + stevia | 81 [77 - 85] | 80 [76 - 84] | 80 [76 - 85] |
|  | Water | 79 [75 - 84] | 79 [75 - 83] | 83 [79 - 87] |
| **Nacc<sup>1</sup>** | Sucrose | 107 [104 - 110] | 107 [104 - 110] | 107 [104 - 110] |
|  | Sucralose | 106 [103 - 109] | 108 [105 - 111] | 109 [106 - 112] |
|  | Stevia | 106 [103 - 109] | 111 [108 - 114] | 109 [106 - 112] |
|  | Monk fruit | 106 [103 - 109] | 106 [103 - 109] | 108 [105 - 111] |
|  | Allulose + stevia | 106 [103 - 109] | 109 [106 - 112] | 108 [105 - 111] |
|  | Water | 106 [103 - 109] | 105 [102 - 108] | 111 [108 - 114] |
| **Substantia nigra** | Sucrose | 59 [56 - 61] | 59 [56 - 61] | 59 [57 - 62] |
|  | Sucralose | 58 [56 - 61] | 60 [57 - 62] | 59 [57 - 62] |
|  | Stevia | 58 [56 - 60] | 60 [58 - 63] | 59 [56 - 61] |
|  | Monk fruit | 58 [56 - 61] | 60 [58 - 62] | 60 [58 - 63] |
|  | Allulose + stevia | 59 [56 - 61] | 58 [56 - 61] | 59 [56 - 61] |
|  | Water | 58 [56 - 61] | 57 [54 - 59] | 59 [57 - 62] |
| **VTA<sup>1</sup>** | Sucrose | 73 [68 - 78] | 75 [71 - 80] | 67 [63 - 72] |
|  | Sucralose | 72 [67 - 77] | 77 [72 - 82] | 79 [74 - 84] |
|  | Stevia | 73 [68 - 78] | 69 [64 - 74] | 72 [67 - 77] |
|  | Monk fruit | 72 [67 - 77] | 74 [69 - 79] | 77 [72 - 82] |
|  | Allulose + stevia | 72 [67 - 77] | 71 [66 - 75] | 73 [68 - 78] |
|  | Water | 73 [68 - 78] | 69 [64 - 73] | 75 [71 - 80] |

<sup>1</sup>Nacc, nucleus accumbens; ROI, region of interest; VTA, ventral tegmental area

For the substantia nigra, there was no effect of treatment (P=0.75) and timepoint (P=0.47) and no interaction between treatment and timepoint (P=0.78). Plots for these ROIs are shown in **Supplementary Figure 4**.

For the VTA (see **Figure 2**), there was no effect of treatment (P=0.28) and timepoint (P=0.40). There was a interaction between treatment and timepoint (P=0.034). At T=5 min, CBF in the VTA tended to be higher for Sucralose compared to Stevia (difference 7.8±3.10, P_FDR_=0.091) and Water (difference 8.26±3.12, P_FDR_=0.091). Compared to Sucrose, VTA CBF at T=30 min was higher for Sucralose (difference 11.29±3.10, P_FDR_=0.004), Monk fruit (difference 9.46±3.12, P_FDR_=0.020) and Water (difference 8.00±3.09, P_FDR_=0.0499), driven by a decrease for Sucrose and increases for Sucralose, Monk fruit and water compared to baseline (**Figure 2A**). In addition, for Sucrose, the VTA CBF change from baseline differed between T=5 and T=30 (7.92±3.09, P_FDR_=0.032), with a CBF increase at T=5 followed by a decrease in CBF at T=30 **(Figure 2B)**. The CBF change at T=30 differed between Sucrose and Sucralose, with a decrease for Sucrose and an increase for Sucralose (difference 12.05±3.12, P_FDR_=0.018).

**Figure 2.**
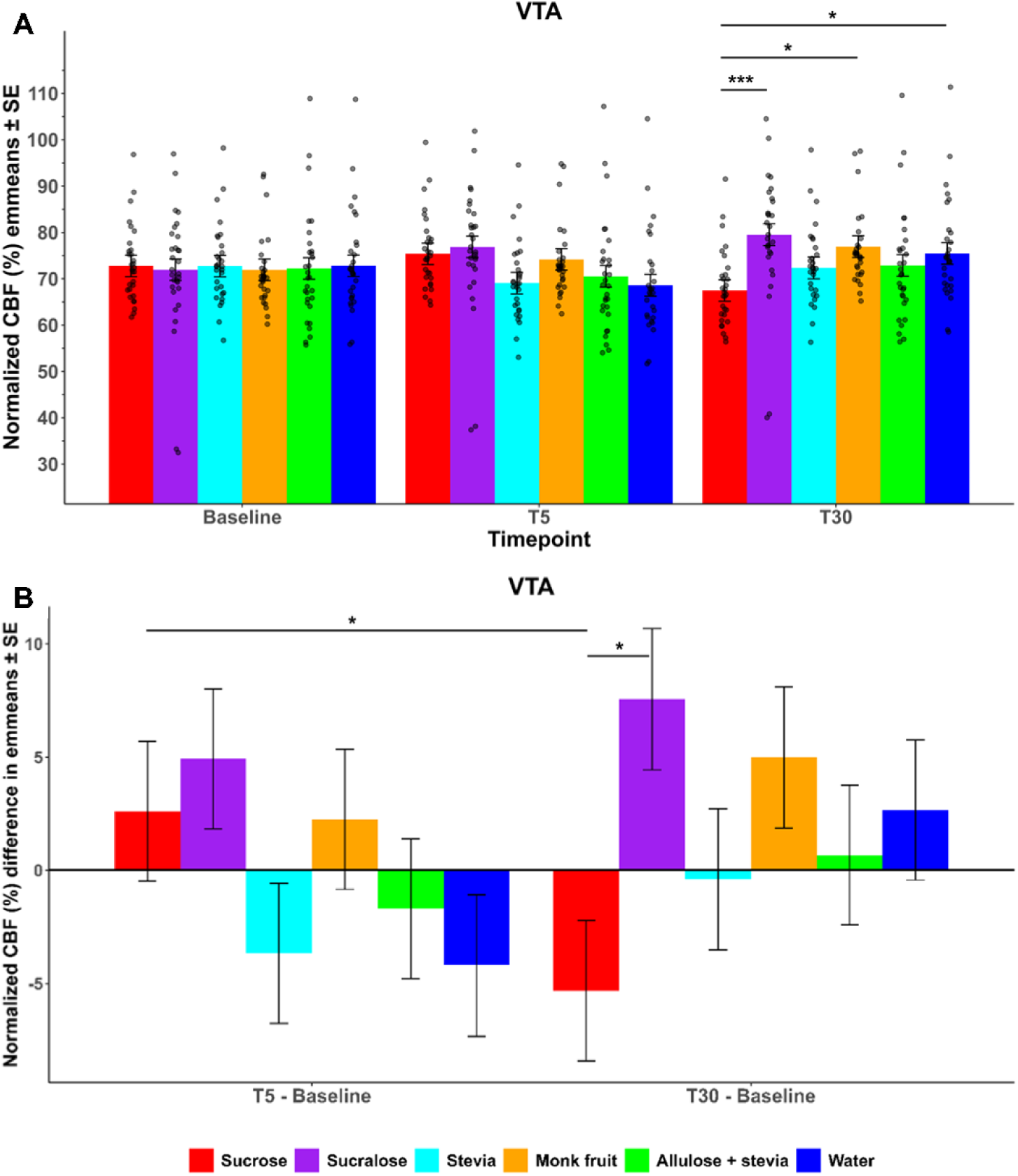
Estimated marginal mean normalized CBF (% of global CBF) ± SE and changes from baseline for the VTA, based on the linear mixed model with baseline and sex as covariates. Data points represent the predicted values. VTA: At T=30 min CBF is lower for Sucrose compared to Sucralose (P_FDR_=0.004), Monk fruit (P_FDR_=0.020) and Water (P_FDR_=0.0499). For Sucrose, the change at T5 differs from that at T30 (P_FDR_=0.032) and the change at T30 differs between Sucrose and Sucralose (P_FDR_=0.018). * P_FDR_ < 0.05; ** P_FDR_ < 0.01; * P_FDR_ < 0.005.

### Exploratory whole-brain analysis (LNCS versus Sucrose)

Results are shown in **Table 2** and **Figure 3**. Compared to baseline: there was an increase in CBF across conditions in the insula, putamen and anterior cingulate cortex at T=5 min (P_FWE_< 0.05 cluster level corrected). This effect was still evident, albeit less pronounced at T=30 min. Furthermore, at T=5 min CBF was decreased in the cuneus and fusiform gyrus. At T=30 min, superior temporal gyrus CBF was decreased (for all results: P_FWE_< 0.05 cluster level corrected). There were two treatment by timepoint interactions for LNCS and sucrose (P_FWE_<0.05, small volume corrected). In the left amygdala, the change in CBF from T=5 to T=30 differed between Sucrose (decrease) and Allulose + stevia (increase). Similarly, in the left putamen, the change in CBF from baseline to T=30 differed between Sucrose (decrease) and Stevia (increase).

**Figure 3.**
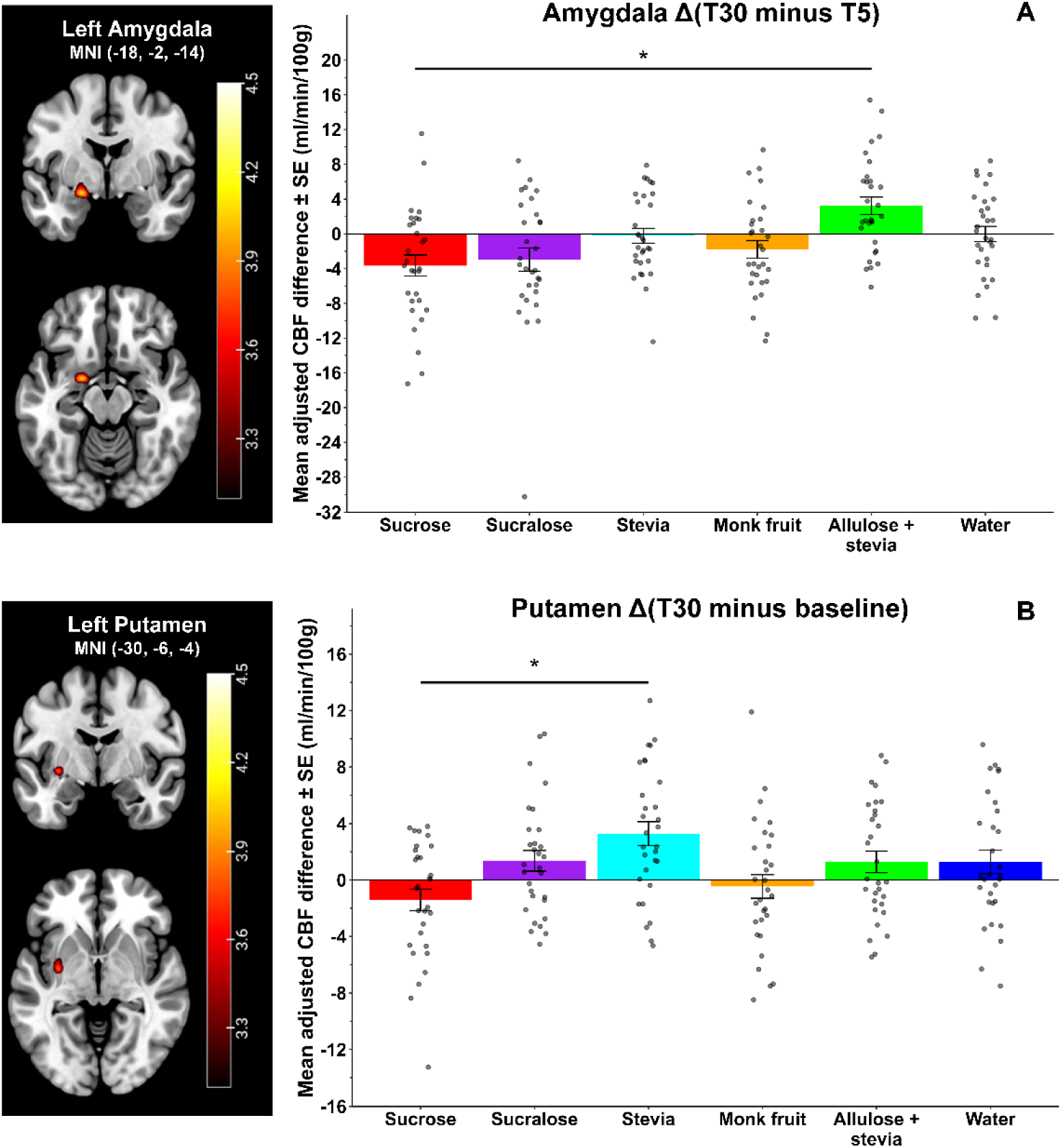
Mean ± SE adjusted CBF differences based on the whole-brain analysis for the clusters in the left amygdala. (A) and left putamen (B) showing a significant time by treatment interaction. For this visualisation the differences for all treatments are shown. BL, Baseline. The colored bar shows the color-coding of the T-values for each voxel which are superimposed on an anatomical brain MRI (MNI152 template). * Interaction P_FWE_ < 0.05, small volume corrected.

**Table 2.**
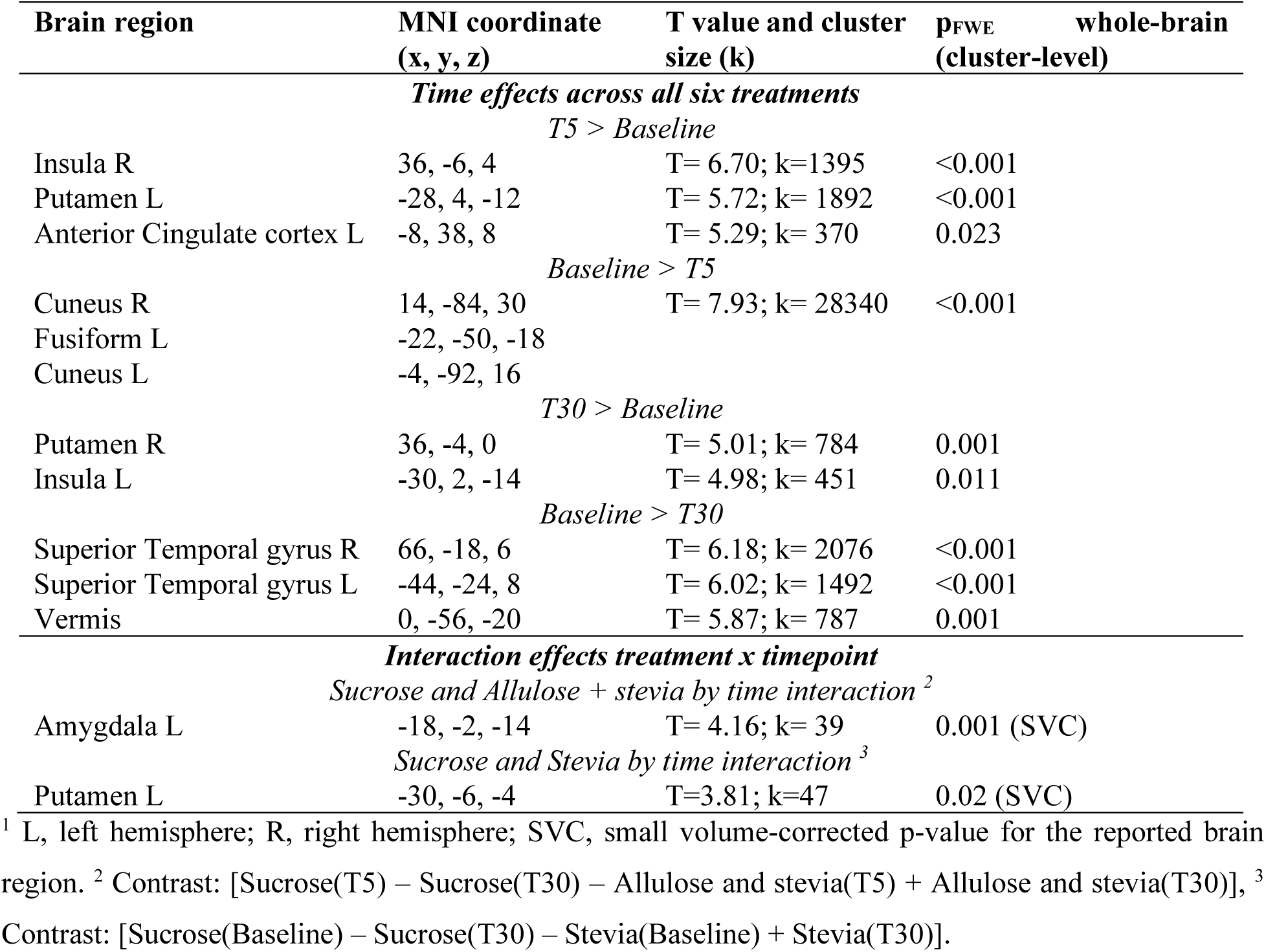
Results of the exploratory whole-brain CBF analysis (flexible factorial model)^1^.

### Glucose and insulin

Results are shown in **Figure 4**. For glucose there was a main effect of time, treatment and a treatment by time interaction (all P<0.001). At baseline and T=5, there were no differences between the drinks. At T=15, T=30 and T=45 min, all drinks differed from Sucrose (all P<0.001). At T=45 min, there tended to be higher glucose for Sucralose compared to Allulose + stevia (Sucralose: 4.81 95% CI 4.66-4.95 versus Allulose + stevia: 4.56 95% CI 4.42-4.71, P=0.10). At T=60 min, Allulose + stevia (P<0.001) and Monk fruit (P=0.013) still had lower glucose than Sucrose; Stevia tended to differ from Sucrose (P=0.087) and Sucralose no longer differed from Sucrose (P=0.208).

**Figure 4.**
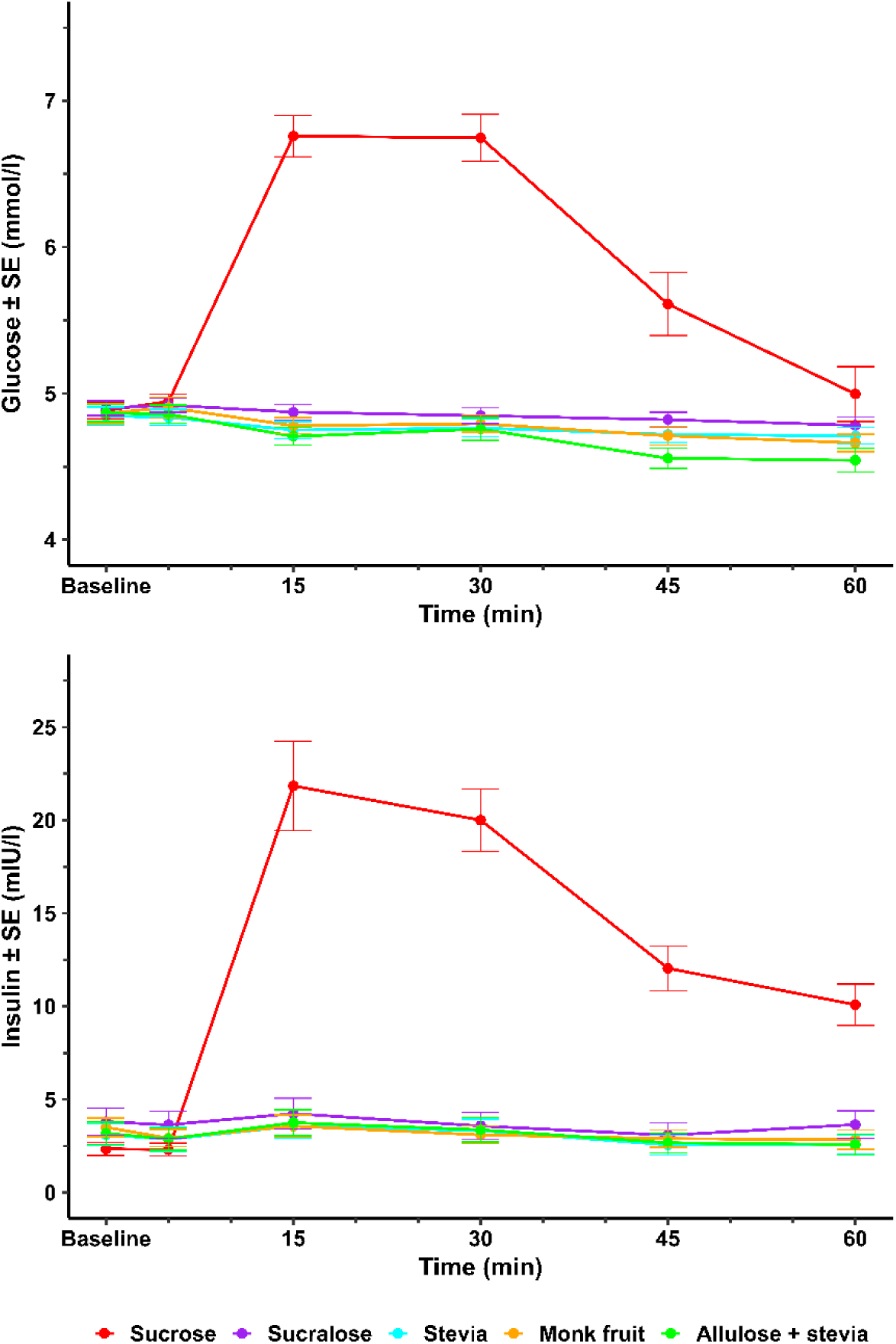
Mean ± SE glucose (top) and insulin (bottom) concentrations over time for all treatments.

For insulin there was a main effect of time, treatment and a treatment by time interaction (all P<0.001). At baseline and T=5, there were no differences between the drinks. For all later timepoints insulin was higher for Sucrose compared to all other drinks (all P<0.001).

### Gastric content volume

Results are shown in **Figure 5**, **Supplementary Figure 6** (unadjusted mean volumes) and **Supplementary Table 2** (differences between treatments per timepoint). There was a main effect of treatment such that gastric content volume (GCV) was higher for Sucrose and Allulose + stevia versus the other drinks, time and a treatment by time interaction (all P<0.001). At baseline there were no differences between the drinks. At T=25, Sucrose had a greater GCV than all other treatments except for Allulose + stevia (Water P_FDR_=<0.001; Sucralose P_FDR_=<0.001; Monk fruit P_FDR_=0.001; Stevia P_FDR_=0.016). Allulose + stevia had a greater GCV than Water (P_FDR_<0.001) and all the other sweet drinks including Sucrose (Sucralose P_FDR_<0.001; Monk fruit P_FDR_<0.001; Stevia P_FDR_<0.001, Sucrose P_FDR_=0.037). At T=45, Sucrose had a larger GCV than Water, Sucralose, Stevia and Monk fruit (all P_FDR_<0.001) and tended to have a greater GCV than Allulose + stevia (P_FDR_=0.053). Allulose + stevia had a greater GCV than Water (P_FDR_=<0.001), Sucralose (P_FDR_=0.012), Stevia (P_FDR_=0.007) and Monk fruit (P_FDR_=0.026).

**Figure 5.**
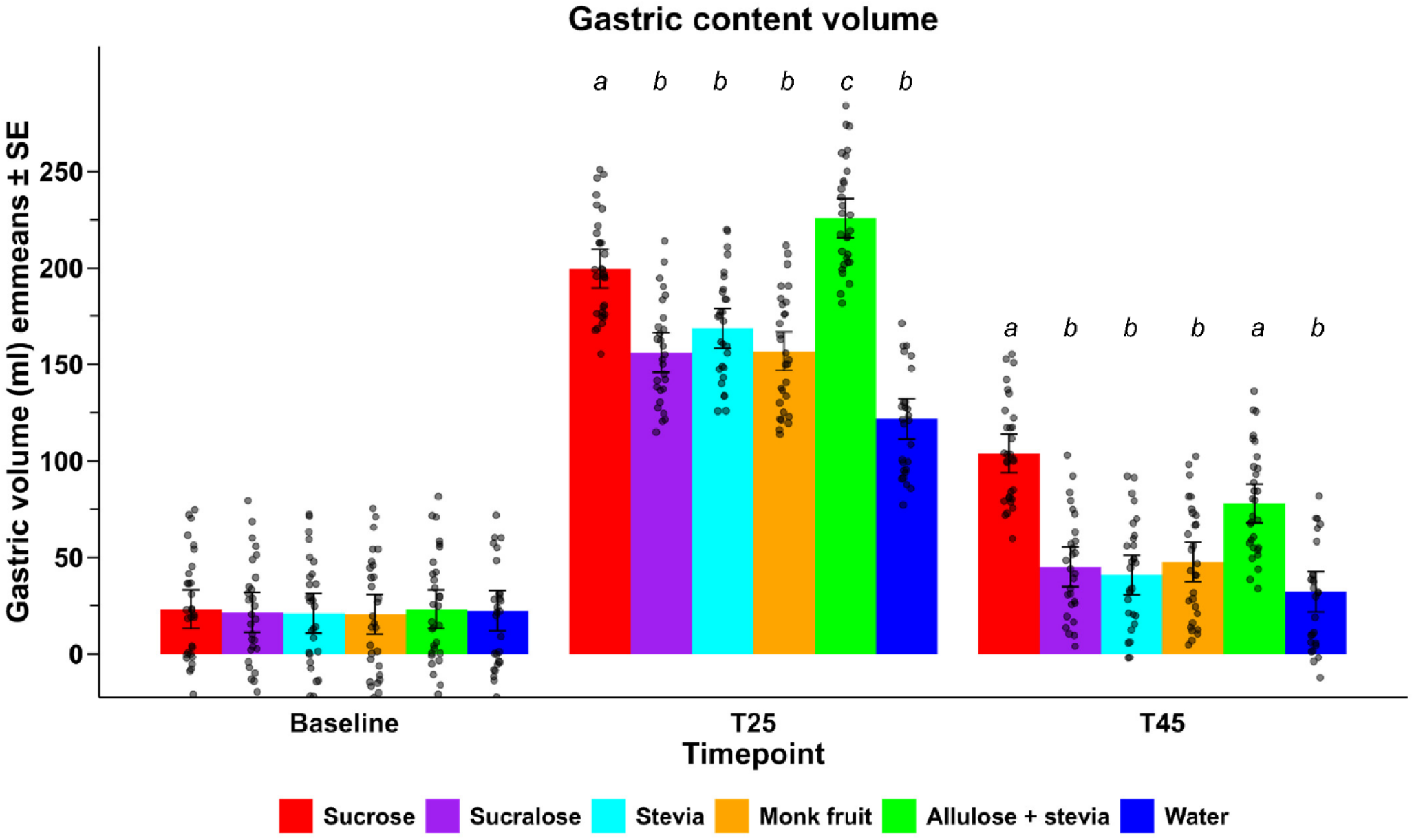
Estimated marginal means (emmeans) ± SE for gastric content volume based on the linear mixed model with baseline and sex as covariates. Data points represent the predicted values. Different letters indicate that treatments differ significantly at a timepoint (P_FDR_<0.05). A plot of the unadjusted means can be found in Supplementary Figure 5.

### Subjective ratings

Data are shown in **Supplementary Figure 7**. For hunger there was a main effect of treatment and time (both P<0.001) but no interaction effect (P=0.999). Hunger was lower for Sucrose versus Water (5.22±1.23, P_FDR_<0.001), Sucrose versus Stevia (3.16±1.21, P_FDR_=0.035), Sucralose versus Water (3.82±1.22, P_FDR_=0.014) and Allulose+stevia versus Water (3.53±1.24, P_FDR_=0.022). It tended to be lower for Monk fruit versus Water (2.89±1.22, P_FDR_=0.053).

For fullness there were treatment (P=0.001) and time (P<0.001) effects but no interaction effect (P=0.410). However, there were no significant treatment differences post hoc.

For desire to eat, there was a main effect of treatment and time (both P<0.001) but no interaction effect (P=0.996). Desire to eat was lower for all sweeteners versus Water (Sucrose 5.15±1.29, P_FDR_<0.001; Sucralose 5.17±1.28, P_FDR_<0.001; Stevia 4.24±1.29, P_FDR_=0.005; Monk fruit 3.98±1.28, P_FDR_=0.007; Allulose+stevia 3.70±1.29, P_FDR_=0.013).

Thirst declined after consumption but did not differ between the treatments (P=0.232).

Bloating ratings were very low, around 20, and showed no differences between treatments (all P < 0.27). Wellbeing was scored around 70 and did not differ between treatments (P=0.448).

## Discussion

Drinks with LNCS do not contribute to energy intake while still providing the hedonic experience of sweet taste. LNCS can have differential effects on brain areas involved in food reward and the regulation of food intake compared to sugars as shown in response to simple solutions with high doses^5,6,24,27,32,52^. We examined changes in cerebral blood flow in response to the ingestion of flavored waters sweetened using a lower dose of 25 g of the sugar sucrose or LNCS and water. In addition, changes in gastric content volume, insulin and glucose levels (for all sweet treatments) and appetite were measured.

While no significant differences in hypothalamus blood flow and appetite rating were observed, LNCS ingestion resulted in distinct brain and gastric responses compared to 25 g of sucrose. Drinks containing allulose + stevia, or stevia alone, altered activity in reward-related brain regions. The allulose + stevia drink also delayed gastric emptying to a degree comparable to the sucrose drink, while only that drink increased glucose and insulin.

### Brain responses

In the hypothalamus, the primary region of interest, no differential treatment effects were found. This contrasts with previous studies reporting differences between glucose or fructose ingestion and water on hypothalamus activity using functional MRI (either CBF^29,31^ or BOLD signal^27,30^). Unlike most previous studies, we used the disaccharide sucrose (i.e., glucose + fructose), which is commonly used in beverages in Western Europe. Page et al.^29^ reported a decline in hypothalamus CBF after ingestion of 75 g glucose, but not fructose, dissolved in 300 ml flavored water. In a recent study involving a relatively large sample (n=75) with a broad BMI range, the effect of a 300 ml drink containing 75 g of sucrose on hypothalamus blood flow did not differ from that of water^32^. Specifically, in individuals with healthy weight (n=26) the decline in lateral hypothalamic blood flow was greater after 75 g sucrose compared to sucralose but did not differ from water^32^. Van Opstal et al.^27^ found a more delayed and smaller decrease in hypothalamus activity after ingestion of 50 g fructose and sucrose compared with glucose ingestion and attribute this to the slower metabolization of fructose. Intra-gastric loads of the sugar alcohol xylitol (50 g in 300 ml water), but not erythritol (75 g), increased CBF in the hypothalamus, whereas 75 g glucose in 300 ml water had the opposite effect^31^. In line with these findings, dose-dependent BOLD signal decreases were reported in parts of the hypothalamus after ingestion of 25 and 75 g glucose in 300 ml water^53,54^. Although this was considered in our sample size estimation, the dose of 25 g sucrose in 500 ml may have been too low to elicit a sustained reduction in hypothalamus activity, as reflected by lower CBF. The overall effect of sucrose on the hypothalamus might be small, since fructose tended to increase hypothalamus CBF^29^, potentially counteracting any glucose-related decrease. In the VTA, a core hub for reward processing, CBF was higher at 30 min for Sucralose, Monk fruit and Water compared to Sucrose, with the largest CBF increase in response to Sucralose. This aligns with the finding that VTA activity decreased with glucose but increased similarly for water and sucralose (matched to 50 g of glucose in 300 ml water) up to 22 min after ingestion^27^. This was proposed to be due to the absence of energy and possibly a reflection of lower satiety^27^. Likewise, in our study, a higher VTA response was identified at the later time point but not shortly after ingestion, pointing to the possible influence of satiety signaling on VTA activity rather than an acute effect of the sweet taste. The drinks with allulose+stevia and stevia did not show different CBF in the VTA at 30 min, indicating differential responses based on the LNCS type. The VTA plays a key role in dopamine signaling in the mesolimbic reward pathway and integrates homeostatic and reward signals to regulate the intake of palatable foods. Whether the differential effects on VTA activity of some LNCS drinks could affect eating behavior needs to be further investigated.

While Stevia and Allulose + stevia did not change CBF in dopaminergic midbrain areas, increased CBF was observed in the left putamen and amygdala for Stevia and Allulose + stevia. This is in line with previous studies showing differential striatal and amygdala activation and functional connectivity in reponse to different LNCS^20,31,52^. Further studies are needed to investigate the effects of different types of LNCS or combinations of sweeteners on reward processing.

In summary, we found differences in the effects of the ingestion of various sweeteners on brain regions involved in reward, motivation, and energy intake regulation in individuals with a healthy weight. However, the hypothalamus, the primary region of interest, did not show the expected differential treatment effects observed in previous studies^24,29,32,52,55^. It should be noted that these studies differed in participants’ body weight ranges, as well as in the dosage and type of sweeteners used, and did not always adjust their CBF analyses for global CBF changes. Most studies used at least 50 g of sugar and in part used flavored water as a control, complicating direct comparisons with the current study. All sweet drinks in this study contained a lemon-lime flavor which could not be used in the water treatment. Thus, comparisons with water could show effects of the flavor next to sweetness effects. However, the data show no indication of flavor effects, which would likely be short-lived.

### Glycemic responses, gastric emptying and appetite

As expected, only Sucrose resulted in increased blood glucose and insulin concentrations. The drink with 25 g allulose + stevia, which is low-caloric because allulose provides 90 – 95 % less energy than sucrose^56,57^, showed a very small reduction in blood glucose, in the absence of any insulin effects. While not deemed physiologically relevant, this observation is in line with studies that examined the glucose-lowering effects of allulose when co-ingested with a sugar^58^.

Despite its low energy content Allulose + stevia slowed gastric emptying compared to Water and the LNCS drinks, similar to Sucrose. That drink was expected to show the slowest gastric emptying because of its caloric content. It can be noted that unlike the other LNCS, allulose was the only bulking sweetener used in a higher quantity, which provides a viscosity and mouthfeel similar to that of sucrose, which also has bulking properties. Previous studies have shown that allulose can induce GLP-1 release, which in turn could inhibit gastric emptying. Teysseire et al.^59^ found GLP-1, CCK and PYY release after intragastric administration of 25 g of allulose in 300 ml tap water. Despite the release of GLP-1 and other appetite-related hormones, they observed no significant effect on gastric emptying as measured with a tracer. When paired with lactisole, a competitive inhibitor of the sweet taste receptor (T1R2/T1R3), similar results were obtained. This is in line with earlier work in which blocking the sweet taste receptors in the gastrointestinal tract with lactisole suppressed appetite-related hormone release but did not affect gastric emptying rate of a 75-g intragastric glucose load in 500 ml water (Gerspach et al., 2011). A key difference with our study is that we had oro-sensory exposure to sweetness, matched to that of 50 g/L sucrose, as well as a direct measurement of gastric content volume. Exposure to sweetness in the absence of energy in the form of a diet soda has been shown to augment the GLP-1 response to subsequent ingestion of a 75-g glucose load^61^(Brown et al., 2009). In another study, blocking oral sweet taste perception with lactisole resulted in somewhat slower gastric emptying of a 15% glucose solution while emptying of an equisweet aspartame solution was unaffected^62^. This suggests that sweet taste alone is not sufficient to affect gastric emptying of a non-caloric sweet liquid and that it may act to delay emptying of a caloric sugar solution. Another factor that can affect gastric emptying is osmolarity. Osmolarity of the chyme is sensed by osmoreceptors in the duodenum and non-isotonic duodenal contents slows down gastric emptying^63–65^. Hyperosmolar duodenal content lowers plasma ghrelin concentrations and augments CCK, PYY and GLP-1 concentrations in healthy individuals^66^. In our study the sucrose drink was hypotonic (146 mOsm/L) and Allulose + stevia was isotonic (278 mOsm/L). Thus, the lower gastric emptying rate of Allulose + stevia compared to Water (hypotonic) is likely not due to an osmolarity effect. Taken together, this suggests that the slower gastric emptying of the allulose + stevia drink may have been driven by allulose-induced GLP-1 release. However, this slower gastric emptying did not result in a greater reduction in desire to eat than seen for the other sweet drinks; all sweet drinks were associated with modestly lower desire to eat scores (∼5 points/100). For hunger the decline was smaller and only significant for Sucrose, Sucralose and Allulose + stevia.

### Conclusion

In conclusion, various LNCS-flavored waters generally did not alter regional cerebral blood flow or gastrointestinal responses compared with water ingestion in healthy normal-weight adults, although some differences in reward-related brain areas were noted compared with a 25-g sucrose drink. Some differential effects related to allulose + stevia and stevia ingestion were observed in brain regions involved in the regulation of food intake, warranting further exploration. Allulose + stevia delayed gastric emptying similar to an equisweet sucrose drink, consistent with its ability to induce GLP-1 release. Overall, this suggests that certain non-nutritive sweeteners can have specific neural and gastrointestinal effects despite their minimal energy content. However, different LNCS and water did not differ in their acute glycemic effects and there were no differential effects on appetite of the different sweeteners. Further exploration of the specific neural and physiological effects of stevia extract and allulose is warranted.

## Data Availability

All data underlying the analyses reported in the manuscript are available upon reasonable request to the authors.

## Acknowledgements

We thank Fleur van Duijnhoven, Hester Boekhoud and Xinyang Li for their help with data collection and determining gastric content volumes. The use of the 3T MRI facility has been made possible by Wageningen University Shared Research Facilities.

Author contributions are as follows: PAMS and DR designed the research; EO, PS and SM conducted the research; PS, SK and RV analyzed data; HP, PS, RV and SK interpreted the results; PS, RV and SK wrote the paper. PS and SK had primary responsibility for final content. All authors provided feedback on the draft manuscript and read and approved the final manuscript.

## Abbreviations

ASL: Arterial spin labeling
BOLD: Blood oxygen level-dependent
CBF: Cerebral blood flow
FDR: False discovery rate
FWE: Family-wise error
fMRI: Functional MRI
GCV: Gastric content volume
LNCS: Low-no-calorie sweeteners
MRI: Magnetic Resonance Imaging
Nacc: Nucleus accumbens
pCASL: Pseudo-continuous arterial spin labeling
ROI: Region of interest
rsfMRI: Resting state functional MRI
SN: Substantia nigra
VTA: Ventral tegmental area

