## Supplementary Materials for "Brain and physiological responses to flavored waters with different sweeteners: a randomized cross-over study in healthy young adults"

**Contents**
- Participant flow chart
- Drink composition and perception
- ROI masks and CBF plots
- Gastric content volume
- Subjective ratings

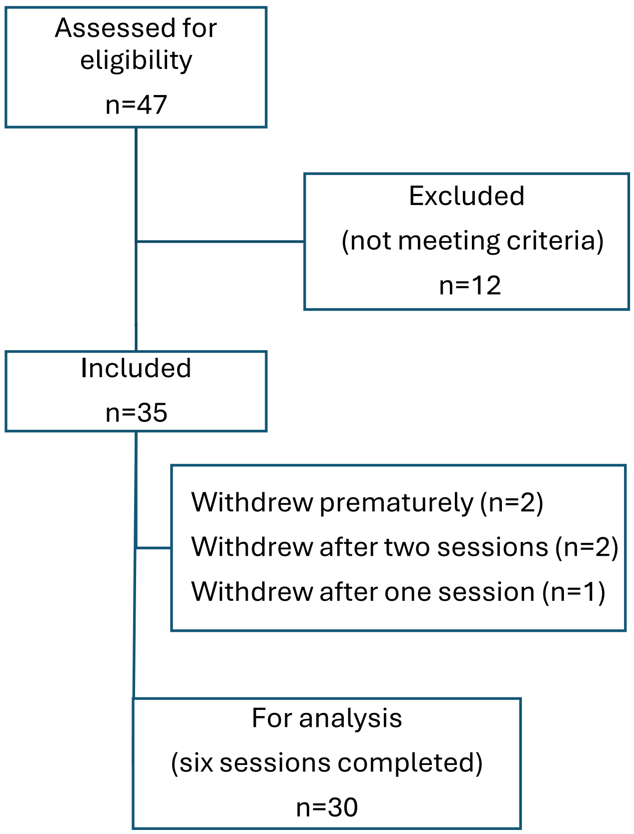


**Supplementary Figure 1**. Participant flow chart.

Drink composition and perception

**Supplementary Table 1.** Drink composition.

| | FORMULATIONS | | | | | | | | | | | |
|---|---|---|---|---|---|---|---|---|---|---|---|---|
| **LEMON-LIME FLAVOURED WATER** | **Sucrose Reference** | | **Sucralose** | | **Tasteva** | | **Monk Fruit M50 (PUREFFRUIT Select) extract** | | **Allulose 3.5 SEV + Tasteva 1.5 SEV** | | **Negative control MINERAL WATER** | |
| **Ingredient name** | **% (cp)** | **g/500 mL** | **% (cp)** | **g/500 mL** | **% (cp)** | **g/500 mL** | **% (cp)** | **g/500 mL** | **% (cp)** | **g/500 mL** | **% (cp)** | **g/500 mL** |
| Flavour Citron vert - 58507 Mane | 0.035 | 0.175 | 0.035 | 0.175 | 0.035 | 0.175 | 0.035 | 0.175 | 0.035 | 0.175 | | |
| Flavour Limette - 10325551 Tagasako | 0.085 | 0.425 | 0.085 | 0.425 | 0.085 | 0.425 | 0.085 | 0.425 | 0.085 | 0.425 | | |
| **Sucrose** | **5.000** | 25.000 | | | | | | | | | | |
| **Sucralose** | | | **0.0083** | 0.0415 | | | | | | | | |
| **Tasteva Stevia** | | | | | **0.0243** | 0.1217 | | | **0.0044** | 0.022 | | |
| **Monk Fruit M50 (PUREFFRUIT Select) extract** | | | | | | | **0.02924** | 0.146 | | | | |
| **Allulose** | | | | | | | | | **5.000** | 25.000 | | |
| Citric Acid VWR | 0.168 | 0.840 | 0.168 | 0.840 | 0.168 | 0.840 | 0.168 | 0.840 | 0.168 | 0.840 | | |
| Preservative: Potasium Sorbate VWR | 0.025 | 0.125 | 0.025 | 0.125 | 0.025 | 0.125 | 0.025 | 0.125 | 0.025 | 0.125 | | |
| Mineral water VOLVIC Danone | 94.687 | 473.435 | 99.679 | 498.394 | 99.663 | 498.313 | 99.658 | 498.289 | 94.683 | 473.413 | 100.000 | 500.000 |
| **TOTAL** | 100.00 | 500.00 | 100.00 | 500.00 | 100.00 | 500.00 | 100.00 | 500.00 | 100.00 | 500.00 | 100.00 | 500.00 |

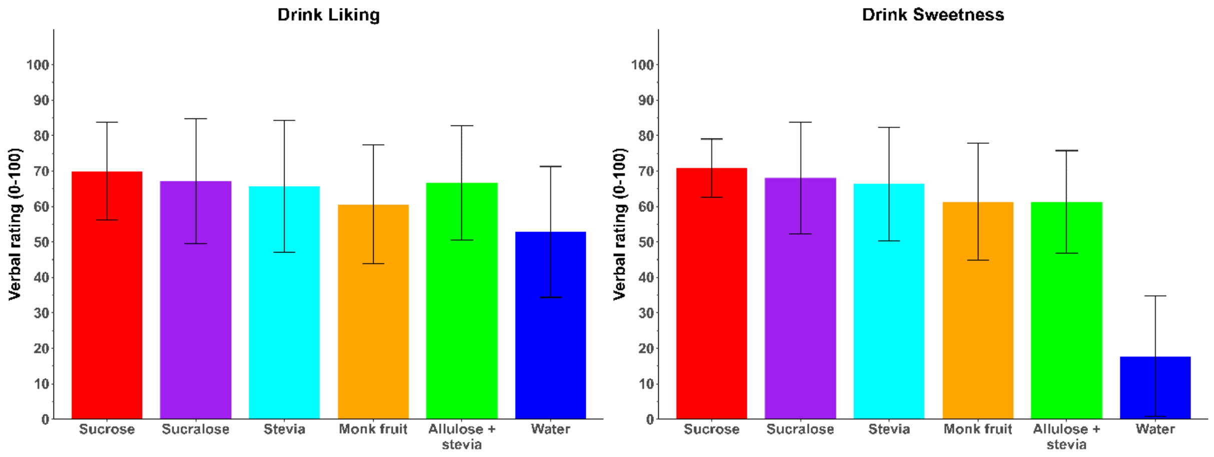


**Supplementary Figure 2**. Mean±SD liking and sweetness ratings for the six treatments. Liking did not differ among the flavored drinks. All drinks except Monk fruit were liked significantly more than Water (all $p<0.05$). Perceived sweetness of all drinks was greater than that of water (all $p<0.001$). Monk fruit and Allulose + stevia tended to be a little bit less sweet than Sucrose (both $p=0.08$) but sweetness did not differ among the flavored drinks (all $p>0.1$, Tukey's HSD posthoc test).

ROI masks and CBF plots

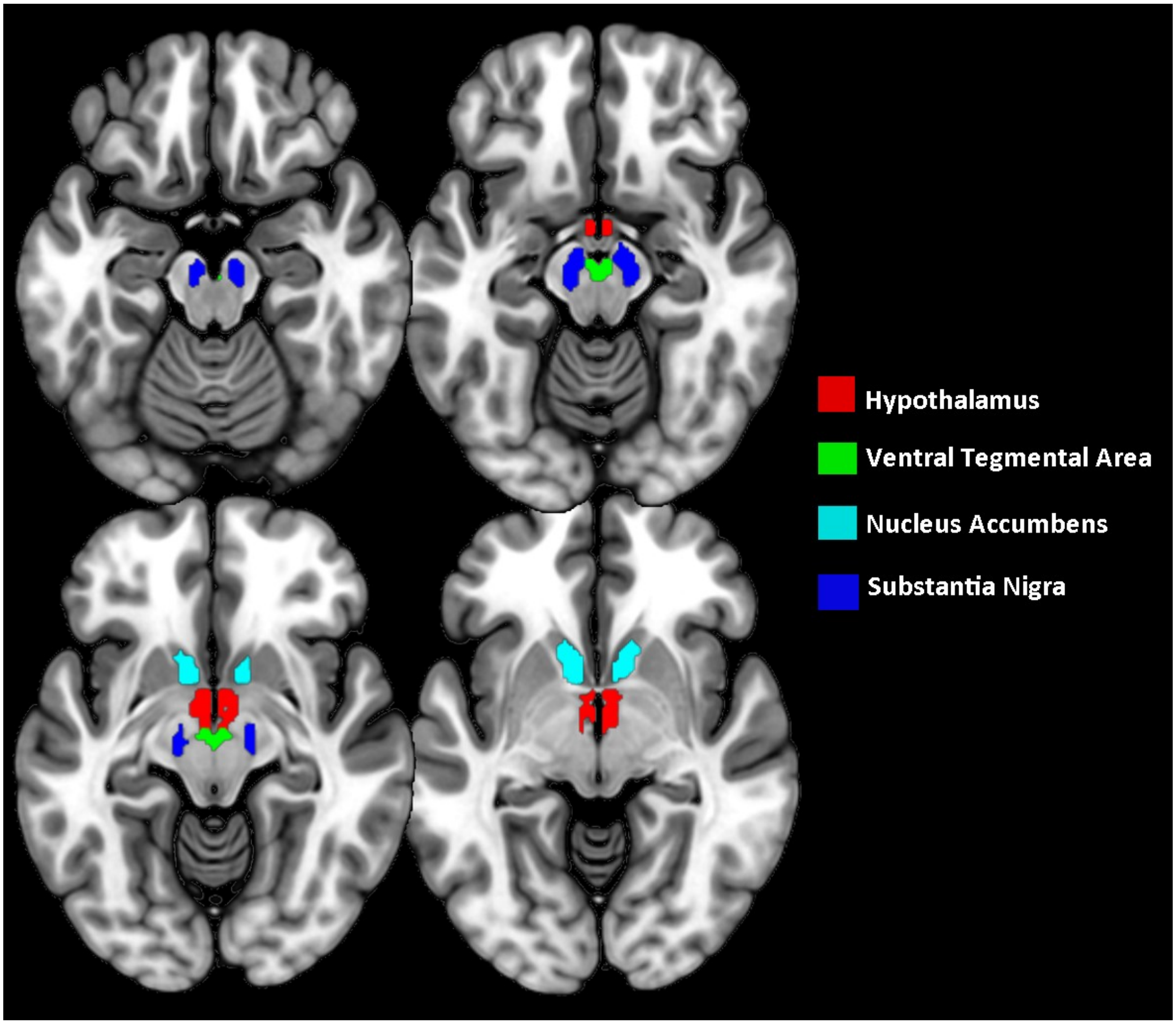


**Supplementary Figure 3**. Illustration of the region of interest masks of the hypothalamus, ventral striatum (Nucleus Accumbens) and dopaminergic midbrain (Ventral Tegmental Area, Substantia Nigra), overlaid on axial crossections.

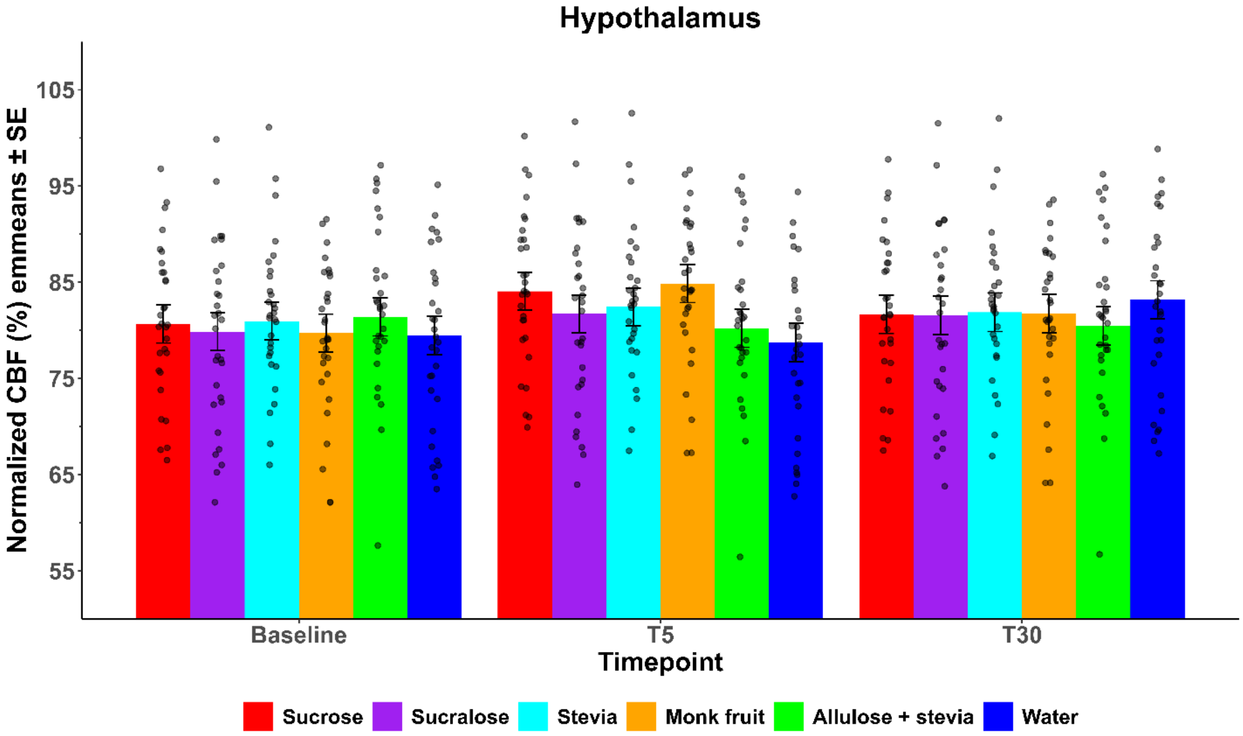
Hypothalamus
Normalized CBF (%) emmeans ± SE
105
95
85
75
65
55
Baseline
T5
T30
Timepoint
Sucrose
Sucralose
Stevia
Monk fruit
Allulose + stevia
Water

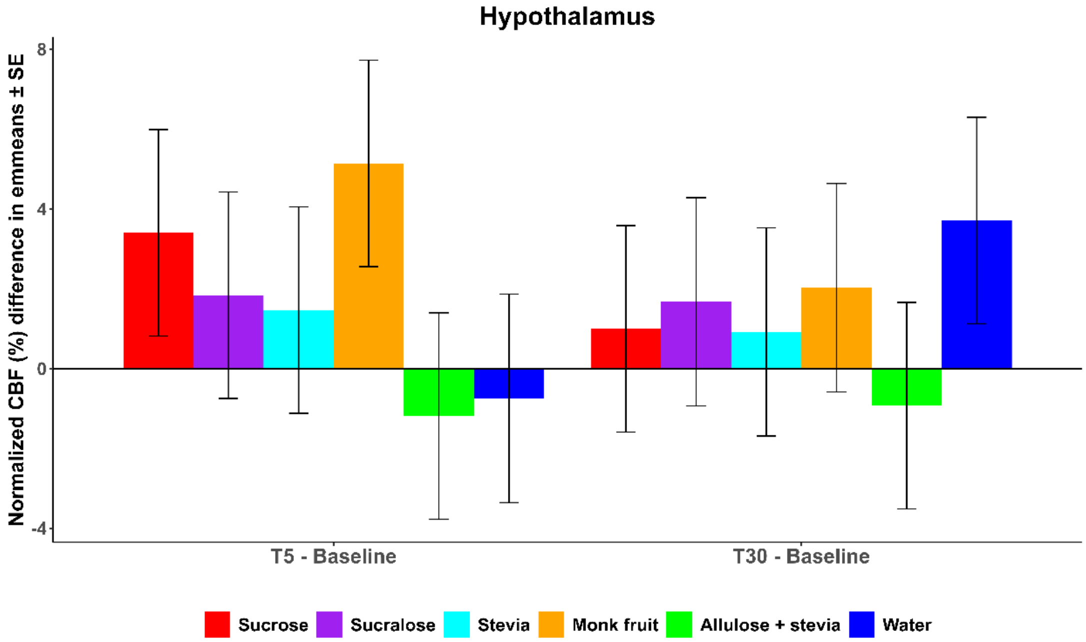
Hypothalamus
Normalized CBF (%) difference in emmeans ± SE
8
4
0
-4
T5 - Baseline
T30 - Baseline
Sucrose
Sucralose
Stevia
Monk fruit
Allulose + stevia
Water

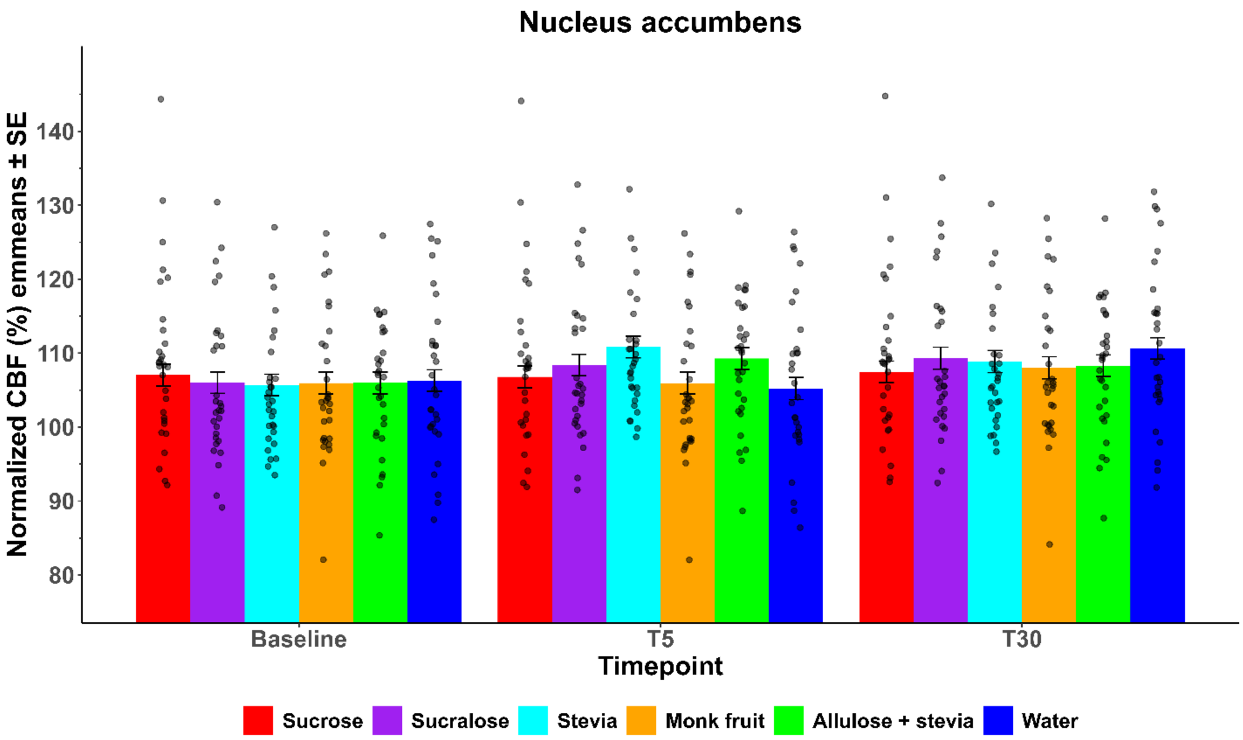
Nucleus accumbens
Normalized CBF (%) emmeans ± SE
140
130
120
110
100
90
80
Baseline
T5
T30
Timepoint
Sucrose
Sucralose
Stevia
Monk fruit
Allulose + stevia
Water

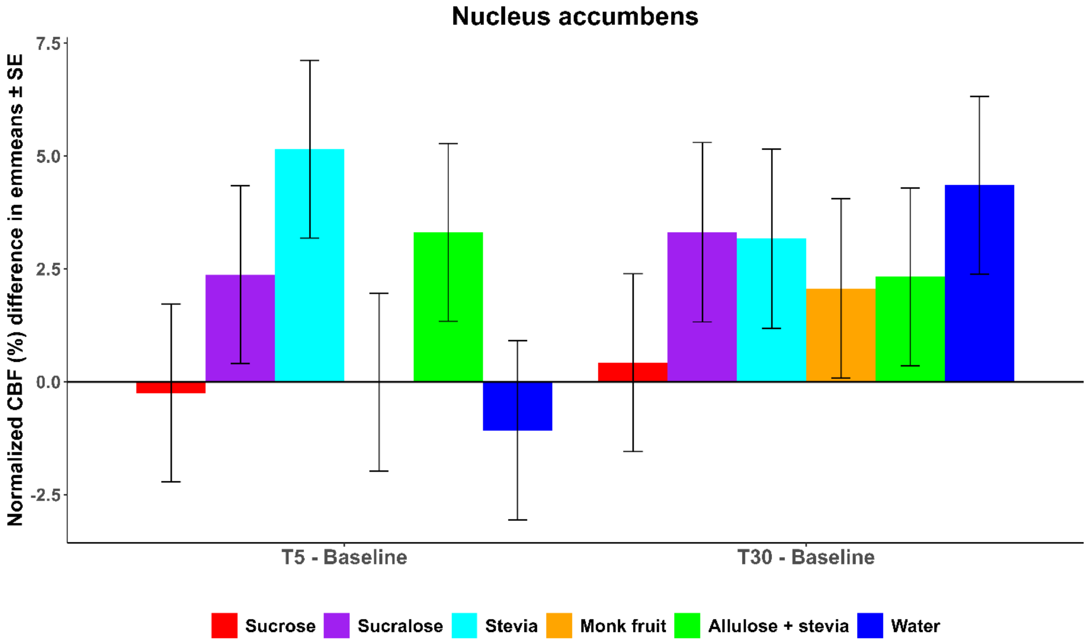
Nucleus accumbens
Normalized CBF (%) difference in emmeans ± SE
7.5
5.0
2.5
0.0
-2.5
T5 - Baseline
T30 - Baseline
Sucrose
Sucralose
Stevia
Monk fruit
Allulose + stevia
Water

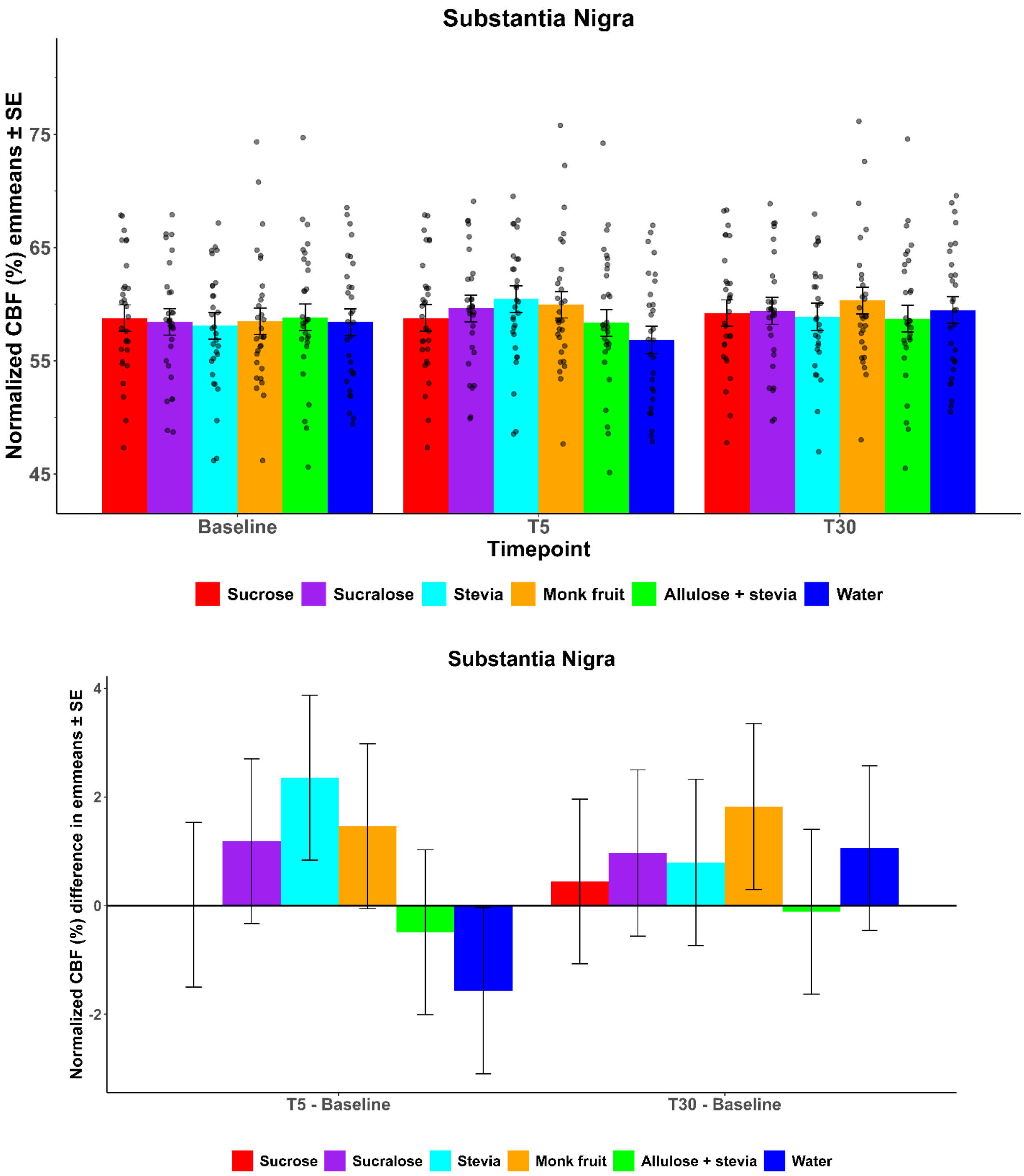


**Supplementary Figure 4**. Estimated marginal mean normalized CBF (% of global CBF) ± SE and changes from baseline, based on the linear mixed model with baseline and sex as covariates for the Hypothalamus, Nucleus Accumbens and Substantia Nigra ROIs.

Gastric content volume

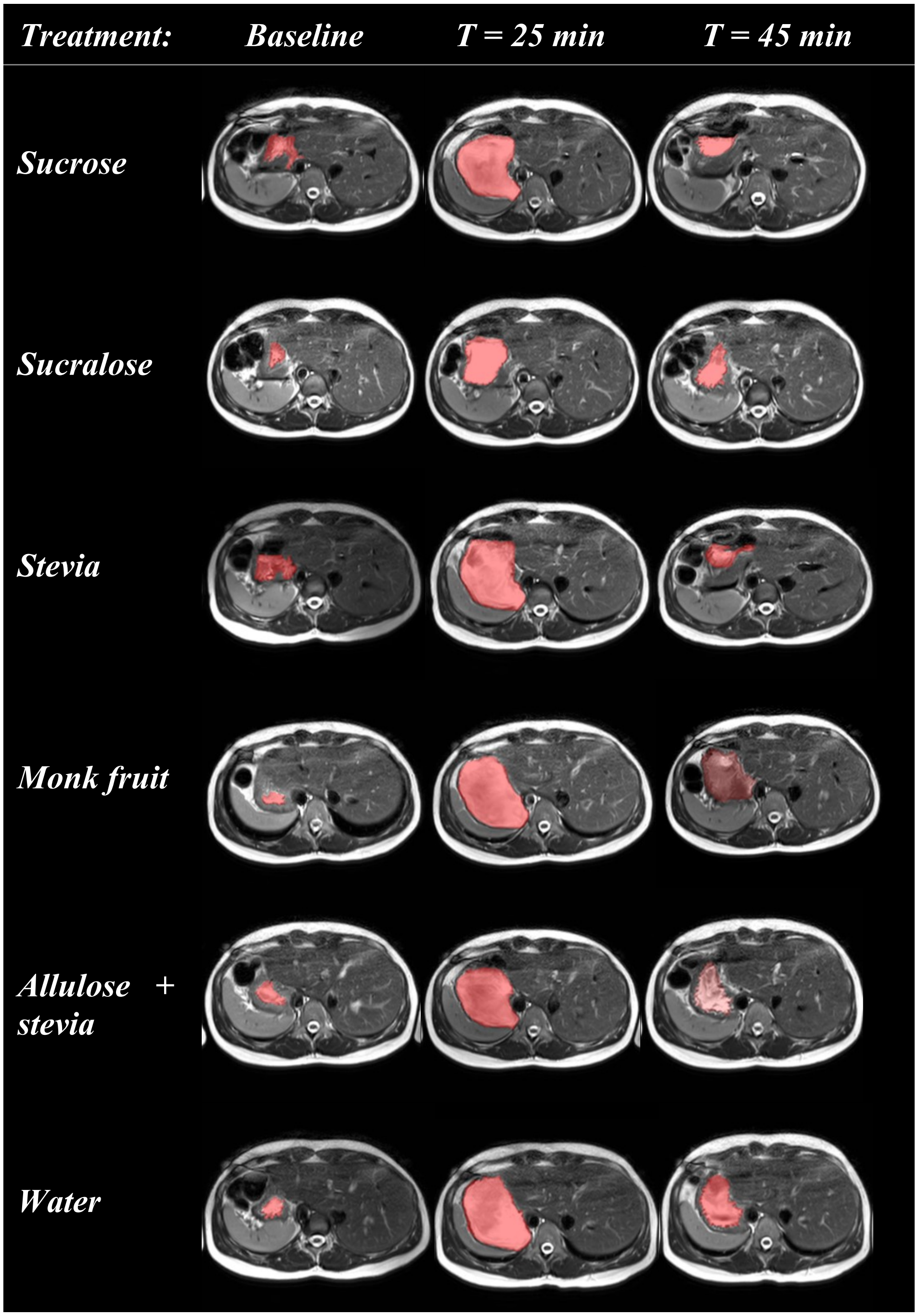


**Supplementary Figure 5**. Illustration of stomach MRI scan time series for all treatments (one participant). Shown is the same axial cross-section. Stomach content is highlighted in red.

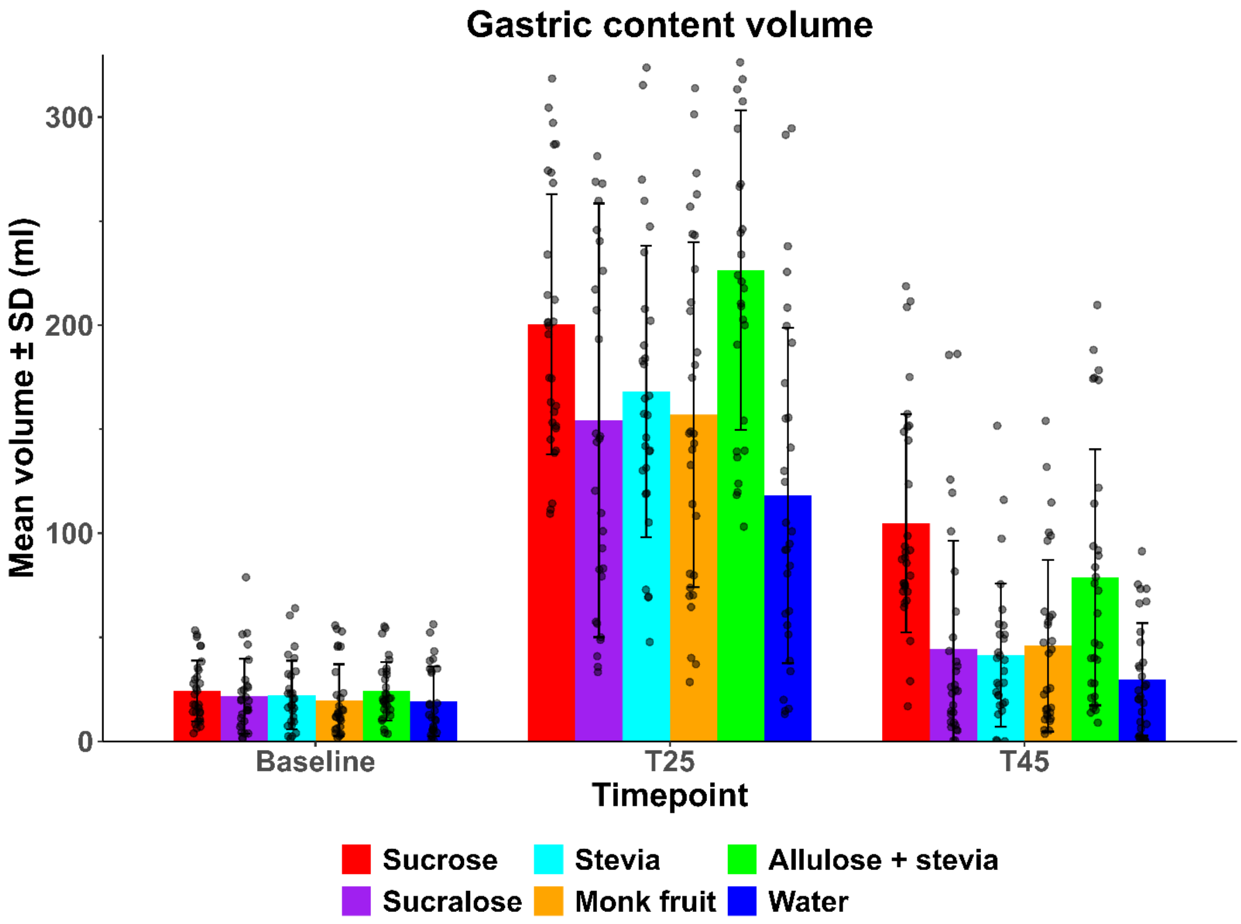


**Supplementary Figure 6**. Mean ± SD gastric content volume for all treatments per timepoint.

**Supplementary Table 2**. Estimated marginal mean differences ± SE of gastric volume (ml) between treatments for each timepoint, based on the linear mixed model with baseline and sex as covariates.

| **Contrast** | **Baseline** | | **T25** | | **T45** | |
|---|---|---|---|---|---|---|
| | **difference** | **$P_{FDR}$** | **difference** | **$P_{FDR}$** | **difference** | **$P_{FDR}$** |
| Sucrose - Sucralose | 1.7 ± 12.2 | 0.9994 | 43.6 ± 12.2 | 0.0009* | 58.9 ± 12.2 | <.0001* |
| Sucrose - Stevia | 2.3 ± 12.2 | 0.9994 | 31.0 ± 12.2 | 0.0158* | 63.1 ± 12.2 | <.0001* |
| Sucrose - Monk fruit | 2.7 ± 12.1 | 0.9994 | 42.8 ± 12.1 | 0.0009* | 56.3 ± 12.1 | <.0001* |
| Sucrose - (Allulose + stevia) | 0.0 ± 12.0 | 0.9994 | -26.2 ± 12.1 | 0.0389* | 26.0 ± 12.0 | 0.0511# |
| Sucrose - Water | 0.9 ± 12.4 | 0.9994 | 77.8 ± 12.4 | <.0001* | 71.7 ± 12.4 | <.0001* |
| Sucralose - Stevia | 0.5 ± 12.5 | 0.9994 | -12.6 ± 12.5 | 0.3619 | 4.2 ± 12.5 | 0.7890 |
| Sucralose - Monk fruit | 1.0 ± 12.3 | 0.9994 | -0.8 ± 12.3 | 0.9498 | -2.6 ± 12.3 | 0.8329 |
| Sucralose - (Allulose + stevia) | -1.7 ± 12.2 | 0.9994 | -69.8 ± 12.3 | <.0001* | -32.9 ± 12.2 | 0.0161* |
| Sucralose - Water | -0.9 ± 12.6 | 0.9994 | 34.3 ± 12.6 | 0.0100* | 12.8 ± 12.6 | 0.4228 |
| Stevia - Monk fruit | 0.5 ± 12.3 | 0.9994 | 11.8 ± 12.3 | 0.3646 | -6.8 ± 12.3 | 0.6717 |
| Stevia - (Allulose + stevia) | -2.3 ± 12.2 | 0.9994 | -57.2 ± 12.3 | <.0001* | -37.0 ± 12.2 | 0.0065* |
| Stevia - Water | -1.4 ± 12.6 | 0.9994 | 46.8 ± 12.6 | 0.0006* | 8.6 ± 12.6 | 0.6193 |
| Monk fruit - (Allulose + stevia) | -2.7 ± 12.1 | 0.9994 | -69.0 ± 12.2 | <.0001* | -30.3 ± 12.1 | 0.0244* |
| Monk fruit - Water | -1.9 ± 12.5 | 0.9994 | 35.0 ± 12.5 | 0.0085 | 15.4 ± 12.5 | 0.3263 |
| (Allulose + stevia) - Water | 0.8 ± 12.4 | 0.9994 | 104.0 ± 12.5 | <.0001* | 45.6 ± 12.4 | 0.0008* |

Subjective ratings

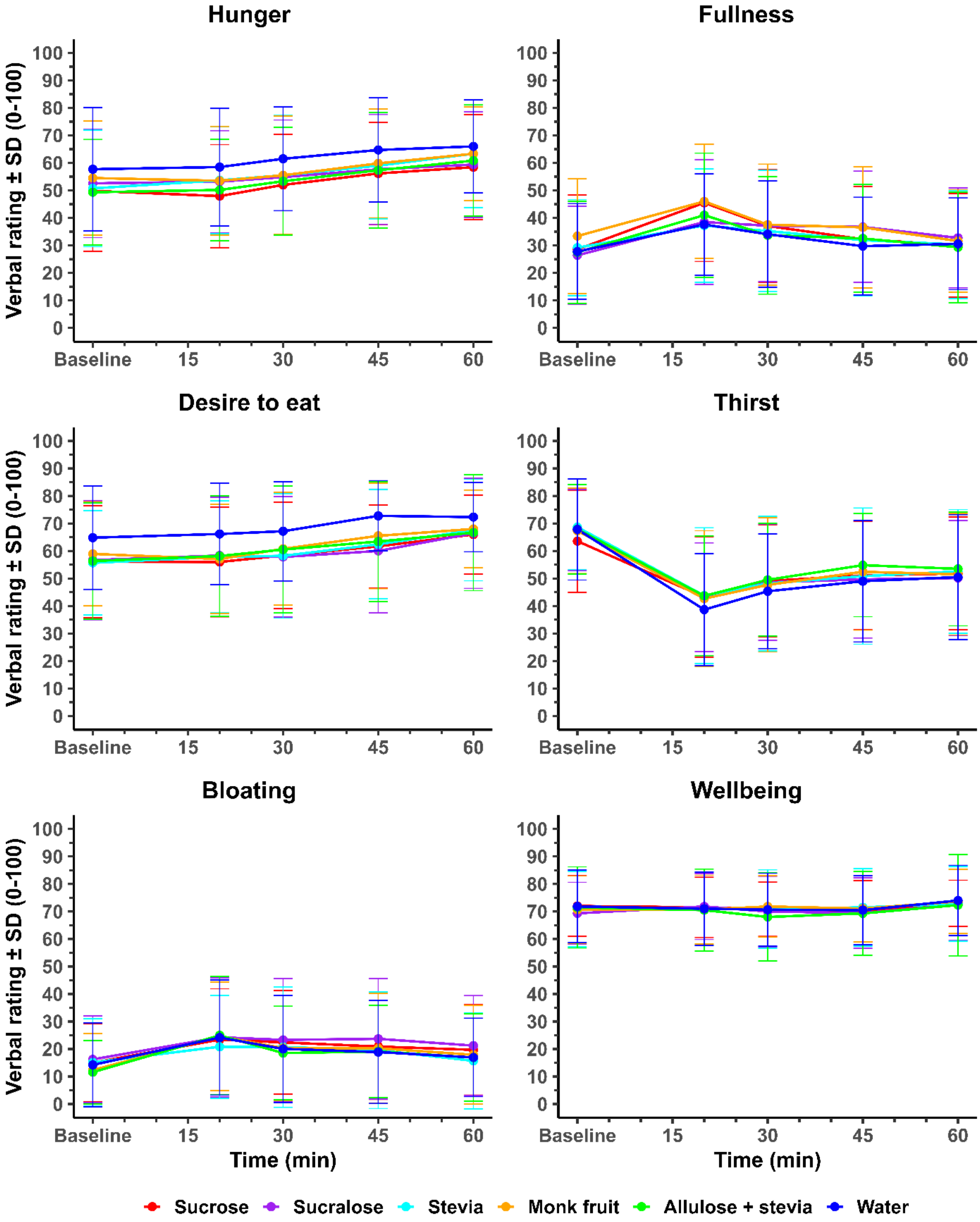


**Supplementary Figure 7**. Mean ± SD subjective ratings over time.
